# From Structural Resources to Latent Protective Capacity: A Bayesian Multilevel Analysis of Flood-Related Depressive Symptoms in Indonesia

**DOI:** 10.64898/2026.08.29.26361712

**Authors:** Suleiman Adamu Yakubu, Seyed Mousavi, Jonathan M. Eden, Judith Kabajulizi, Vasile Palade, Alireza Daneshkhah

**Affiliations:** Centre for Computational Science and Mathematical Modelling, Coventry University, Coventry, UK; Faculty of Mathematics and Data Science, Emirates Aviation University, Dubai, UAE; Centre for Agroecology, Water and Resilience, Coventry University, Coventry, UK; School of Economics, Finance and Accounting, Coventry University, Coventry, UK

**Keywords:** Flooding, depressive symptoms, community resilience, mental health, Bayesian multilevel modelling, latent community protective capacity, disaster risk reduction

## Abstract

Communities exposed to flooding can experience markedly different mental health outcomes, yet conventional resilience indicators capture only part of the social and contextual conditions that may explain this variation. This study develops a multilevel and predictive framework for examining community resilience and depressive symptoms following flood exposure in Indonesia. Data were drawn from 20,303 adults nested within 312 communities in the Indonesia Family Life Survey (IFLS-5). Depressive symptoms were assessed using the 10-item Centre for Epidemiologic Studies Depression Scale (CES-D-10), with Rasch Partial Credit Model calibration used to examine measurement properties. Bayesian multilevel models quantified between-community heterogeneity and assessed how far observable structural resources accounted for this variation. Community resilience was represented through two complementary constructs: structural resilience, based on observable socioeconomic and social-capital resources, and Latent Community Protective Capacity (LCPC), a model-derived proxy for residual contextual variation in depressive-symptom risk. Approximately 6% of variation was attributable to between-community differences, while observable structural resources explained only part of this heterogeneity. Structural resilience and LCPC were weakly correlated (*r* = 0.155). Moderation analyses provided no clear evidence that structural resilience altered the flood–depression association, while LCPC showed a directionally consistent but uncertain buffering pattern. Predictive models incorporating community-level information improved discrimination, with the best-performing model reaching an ROC-AUC of approximately 0.71. The findings suggest that observable resource-based indices provide an incomplete account of community-level mental health vulnerability and that residual contextual measures may provide complementary information, while requiring cautious interpretation and independent validation.

## 1 Introduction

Climate-related disasters are increasing in frequency and intensity, with floods representing the most widespread and recurrent hazard worldwide (Intergovernmental Panel On Climate Change (IPCC), 2023; Centre for Research on the Epidemiology of Disasters (CRED), 2024). Beyond their immediate physical destruction, floods exert substantial and often enduring psychological effects, including elevated risks of depression, anxiety, and post-traumatic stress symptoms (Fernandez et al., 2015; Miller et al., 2024). Yet the mental health consequences of flooding remain poorly integrated into disaster policy and planning. The Sendai Framework for Disaster Risk Reduction 2015–2030 explicitly calls for enhanced recovery schemes that provide psychosocial support and mental health services (United Nations Office for Disaster Risk Reduction, 2015), and comparable commitments appear in national disaster management strategies, but operationalising these commitments confronts a practical obstacle: planners and humanitarian actors lack reliable, scalable tools for identifying *which*communities will carry the heaviest post-flood mental health burden, and therefore where scarce psychosocial resources should be directed. Existing vulnerability indices, built largely from observable material and demographic resources, may not capture the contextual capacities that actually determine how communities absorb and recover from the psychological impact of flooding. Closing this evidence-to-action gap is the central concern of this study.

A recent systematic review and meta-analysis by Yakubu et al. (2026) confirms elevated risks of depression, anxiety, and PTSD across diverse flood-affected populations, highlighting both the consistency of these associations and the role of contextual and socioeconomic modifiers. Miller et al. (2024) similarly identified elevated depression and anxiety prevalence following hurricanes and floods, with effects persisting well beyond the emergency phase and imposing long-term burdens on individuals, households, and health systems. These impacts are particularly pronounced in low- and middle-income countries, where mental health systems remain structurally under-resourced and treatment gaps are wide (Patel et al., 2018), precisely the settings where the ability to target limited resources accurately matters most for policy.

A growing body of research documents substantial community-level variation in post-disaster mental health outcomes, even among populations exposed to broadly comparable hazard intensities (Newnham et al., 2022; Miller et al., 2024; Robin et al., 2020). This mirrors the broader neighbourhood-effects literature in social epidemiology and health geography, which shows that place of residence shapes mental health over and above individual characteristics (Diez Roux and Mair, 2010; Kirkbride et al., 2024). While individual vulnerability factors such as socioeconomic disadvantage and prior health status explain part of this variation, contextual conditions; community socioeconomic composition, social cohesion, institutional responsiveness, and recovery capacity, independently shape post-disaster trajectories (Goldmann and Galea, 2014; Norris et al., 2008), implying measurable between-community heterogeneity within multilevel frameworks.

These observations motivate a community-level resilience lens. Resilience has become a central organising concept in disaster research and policy, yet definitions vary considerably, and this conceptual ambiguity translates into measurement challenges: despite widespread use of resilience indicators, evidence of their predictive validity remains inconsistent (Shiozaki et al., 2024), and contemporary frameworks emphasise a multidimensional capacity to anticipate, absorb, adapt, and recover from shocks (Hutter and Bailey, 2022; Ngai et al., 2022).

Empirical datasets rarely capture all resilience domains simultaneously: large-scale household surveys typically include socioeconomic and social-capital indicators but lack direct measures of infrastructure robustness, governance quality, and institutional preparedness. Constructing resilience solely from additive indices of observable indicators may conflate resources with realised adaptive capacity, while purely latent approaches may lack policy interpretability; composite indices should be grounded in community-specific outcomes (Tariq et al., 2021), and most existing indicator systems lack rigorous outcome-based validation (Shiozaki et al., 2024).

This study adopts such a hybrid conceptualisation by distinguishing between structural resilience and Latent Community Protective Capacity (LCPC). Structural resilience refers to observable socioeconomic and social-capital capacities theorised to enable adaptive responses to disaster exposure. These indicators represent inputs to resilience rather than resilience itself. LCPC, by contrast, is conceptualised as a latent contextual capacity inferred from systematic deviations in mental health outcomes after accounting for individual characteristics and disaster exposure (Norris et al., 2008; Hutter and Bailey, 2022). In multilevel modelling terms, it is approximated through community-level random effects that capture realised protective performance not explained by measured resources (Snijders and Bosker, 2012; Tariq et al., 2021). If structural resilience fully explains between-community variation in depression risk, the estimated community-level variance should attenuate substantially after inclusion of observed resource indicators; persistence of non-trivial residual variance would instead indicate latent contextual capacity beyond observable structural inputs. This logic underpins the variance decomposition strategy adopted here.

Indonesia provides a compelling context. It is among the world’s most disaster-prone countries: floods accounted for over 50% of recorded disaster incidents in recent years (Badan Nasional Penanggulangan Bencana (BNPB), 2024), and exposure is projected to intensify under climate change (Intergovernmental Panel On Climate Change (IPCC), 2023). It is also covered by the Indonesia Family Life Survey (IFLS), a nationally representative longitudinal survey with rich socioeconomic and health modules (Strauss et al., 2016), offering an unusually strong empirical basis for community-level analysis in a low- and middle-income country setting; while not designed to measure disaster resilience, it supports construction of resource-based indicators and inference of latent contextual capacity.

Answering the policy question of where to target post-flood mental health resources requires resolving three prior analytical questions, which this study addresses:

1. How can community resilience be quantified from large-scale household survey data, infrastructure that many disaster-prone countries already maintain, where key resilience domains are only partially observed?
2. To what extent do observable community resources explain between-community heterogeneity in depression risk following flood exposure, and how much protective capacity remains unexplained by the indicators that conventional vulnerability indices rely on?
3. Does incorporating resilience measures improve the identification of communities at elevated mental health risk, in a form that could support the allocation of psychosocial resources?

To answer these questions, we develop a modular analytical framework integrating psychometric calibration, Bayesian multilevel modelling, and complementary machine learning approaches. Bayesian hierarchical models quantify between-community variation while explicitly accounting for uncertainty and clustering; machine learning methods assess predictive performance under flexible specifications. Depression is measured using the CES-D-10, calibrated via a Rasch Partial Credit Model to derive interval-level severity estimates, and additional analyses, including Double Machine Learning (DML), assess robustness. Recent applications of machine learning to flood social vulnerability have largely constructed composite indices from survey data rather than examining community-level resilience as a moderator of flood-related mental health outcomes in population-representative samples (Akindejoye et al., 2025).

By integrating measurement, contextual modelling, variance decomposition, causal robustness checks, and predictive validation within a single analytical architecture, this study makes a contribution positioned at the interface of environmental health science and disaster policy. Analytically, it provides an empirically grounded distinction between structural and latent resilience, addressing measurement challenges identified in prior works (Shiozaki et al., 2024; Tariq et al., 2021; Sedighi et al., 2021), and quantifies how far observable resources explain between-community heterogeneity in depression risk. For policy, it demonstrates that a substantial share of protective capacity is invisible to conventional resource-based vulnerability indices, yet can be recovered from survey data that many disaster-prone states already collect, offering disaster management agencies, health ministries, and humanitarian actors a route to identifying high-need communities that current targeting tools would miss. Although the analysis focuses on Indonesia, the framework is designed to transfer to other low- and middle-income settings where comparable household survey data exist, and to inform the design of analogous tools in higher-income contexts, contributing to the methodological infrastructure for population-based mental health surveillance and resource allocation under escalating climate-related hazard exposure. We return to these policy implications, and their limits, in Section 5.

## 2 Data and Methods

### 2.1 Study Design and Data Source

This study employs a cross-sectional, population-representative design using data from the fifth wave of the Indonesia Family Life Survey (IFLS-5), conducted in 2014-2015. The IFLS is a longitudinal household survey covering 13 of Indonesia’s 27 provinces at baseline (1993), representing approximately 83% of the national population (Strauss et al., 2016). IFLS-5 is the most recent wave and constitutes a large-scale nationally representative dataset, encompassing socioeconomic, demographic, and health information at the individual, household, and community levels. The analytical sample in the current study is restricted to adults aged 15 years and older with valid responses on the depression screening instrument, yielding a working sample of approximately 20,300 individuals nested within 312 original enumeration area (EA) communities. The survey employs a stratified, multi-stage probability sampling design. The primary sampling units are EAs defined at the 1993 baseline; subsequent waves tracked both original households and split-offs (i.e. subclusters of individuals that moved out of the original EAs). Communities are identified by a four-digit EA identifier (`commid_EA`), which forms the primary clustering unit in all multilevel analyses. Analyses are restricted to individuals residing in original IFLS communities, defined as those whose EA code matches a four-digit numeric pattern consistent with the 1993 sampling frame, to preserve the integrity of the community-level clustering structure required for multilevel modelling.

### 2.2 Measures

#### 2.2.1 Outcome: Depressive Symptomatology

Depression was assessed using the Centre for Epidemiologic Studies Depression Scale-10 (CES-D-10), a widely validated brief screening instrument for depressive symptoms in community samples (Andresen et al., 1994). Items were recoded from the original IFLS response scale to the conventional 0–3 metric, two positive-affect items were reverse-scored, and respondents answering fewer than seven of the ten items were excluded. Person-level severity was calibrated under a Rasch Partial Credit Model (Masters, 1982; Mair et al., 2025), yielding an interval-level latent severity score (*θ*) with more robust measurement properties than a simple sum score; full item processing, estimator choices (WLE with EAP fallback), and calibration details are provided in the Appendices (Section A.9).

The primary binary outcome, high depression (`high_depression`), was defined as a raw CES-D-10 sum score ≥ 10, consistent with established cutoffs in community and clinical populations (Andresen et al., 1994; Björgvinsson et al., 2013); a calibrated latent-scale threshold was derived as the median *θ* among respondents scoring exactly 10. This binary outcome was used in all multilevel logistic regression and machine learning models, with the continuous *θ* score retained for threshold sensitivity analyses.

#### 2.2.2 Flood Exposure and Disaster Risk

Household-level flood exposure (flood_exposure) was derived from IFLS-5 disaster modules and defined as a binary indicator of whether the household experienced flooding. This exposure was assigned to all individuals within the same household. Additional disaster-related variables were used to capture frequency, magnitude of the loss and characteristics of the event to construct a larger disaster risk measure. Further details on variable construction are provided in the Appendices (Sections A.2 and A.3).

A composite disaster risk score (`disaster_risk_score`) was constructed as a weighted sum of standardised z-scores for three dimensions: reported disaster frequency (*z_freq*, weight 0.40), maximum asset loss amount (*z_loss*, weight 0.40), and count of disaster types experienced (*z_types*, weight 0.20). When one or more components were unavailable, weights were redistributed proportionally. This continuous index was used in the Bayesian multilevel model as a covariate and in the variance decomposition models as the primary between-community exposure variable.

#### 2.2.3 Individual-Level Covariates

Individual covariates comprised age (mean-centred and scaled; `age_centered`), sex (recorded in IFLS-5 as male/female; variable labelled gender in figures), marital status (four categories), education level (six ordered categories), a household wealth index constructed by principal component analysis of log-transformed consumption, income, and assets (standardised; `wealth_centered`), poor self-rated health (`poor_srh`), any chronic condition, any Activities of Daily Living (ADL) limitation, and urban–rural location derived from community-level administrative coding. Full construction details, coding schemes, and mappings to IFLS source variables are provided in the Appendices (Section A.4, Tables 6–7).

#### 2.2.4 Community-Level Social Trust

Community-level social trust was measured from IFLS social capital items capturing perceived safety, interpersonal attitudes, and community reliability. Items were harmonised to a common scale, reverse-scored where required, and reduced to a single latent factor via one-factor maximum likelihood factor analysis; standardised individual scores (`trust_z`) were aggregated to household and then community (EA) level (`trust_comm_z`). Item selection, coding, and aggregation details are provided in the Appendices (Section A.7).

### 2.3 Community Resilience Operationalisation

The central theoretical construct examined is community resilience, understood as the capacity of a community to attenuate or absorb the adverse mental health effects of flood exposure. We operationalise this construct through two complementary and conceptually distinct indices, drawing on the adaptive capacities framework of Norris et al. (2008), which has been subsequently validated and extended in the disaster resilience literature (Mayer, 2019). This defines community resilience as a set of networked capacities linking economic development, social capital, information systems, and community competence to population wellness outcomes.

#### 2.3.1 Structural Resilience

Structural resilience captures the pre-existing socioeconomic capacity of a community, computed as a weighted composite of five standardised community-level dimensions: household wealth (0.30), consumption (0.25), educational attainment (0.15), urbanisation (0.10), and social trust (0.20). The weighting reflects a theoretically grounded prioritisation of material resources as primary buffering mechanisms in post-flood recovery, consistent with the Baseline Resilience Indicators for Communities (BRIC) framework (Cutter et al., 2010) and empirical evidence from low- and middle-income disaster settings (Norris et al., 2008; Akindejoye et al., 2025). A theory-driven additive composite was adopted over data-driven alternatives such as principal component analysis to preserve the interpretability of resilience dimensions in subsequent moderation analyses. Construction details, the weighting rationale, and sensitivity analyses (equal-weight and PCA-based validation) are provided in the Appendices (Sections B.1–B.2 and C).

#### 2.3.2 Latent Community Protective Capacity

LCPC was derived empirically from the posterior distribution of the Bayesian multilevel model (described in Section 2.5), representing residual variation in depression risk beyond individual level predictors. The primary LCPC measure (`resilience_re`) was defined as the standardised inverse of the community-level random intercept, such that higher values indicate comparatively lower-than-expected depression risk. This measure should be interpreted as a proxy for latent community-level protective capacity rather than as a direct measurement of resilience. It represents residual contextual variation associated with lower-than-expected depression risk after accounting for observed individual and community characteristics, and may therefore reflect the influence of unmeasured resilience-related processes as well as other omitted contextual factors.

Because `resilience_re` is derived from the posterior mean of the community-level random intercepts of the primary Bayesian model, its subsequent use as a predictor in the machine learning analyses introduces a potential generated regressor concern (Pagan, 1984; Murphy and Topel, 1985): the variable is constructed using the depression outcomes of the same individuals on whom the predictive models are trained, meaning the training data has effectively seen outcome-correlated information when constructing one of its predictors. To address this, a province-blocked grouped cross-validation approximation (`resilience_re_gkf`) is used in place of `resilience_re` for the hybrid ML feature sets. Full technical details are provided in Section 2.8 and in the Appendices (Section C.3). Both resilience indices (structural and LCPC) were merged with the individual-level analysis dataset at the community level prior to moderation analyses. Communities were further categorised into quartiles (`resilience_category`) for descriptive and visualisation purposes.

### 2.4 Analytic Framework Overview

The analytic strategy proceeded in four stages: (1) preparation and merging of individual depression scores, covariates, community identifiers, and disaster exposure; (2) a Bayesian multilevel logistic regression characterising determinants of depression risk and deriving the LCPC index; (3) variance decomposition quantifying the community variance attributable to individual covariates, disaster exposure, and structural resilience; and (4) moderation models testing whether resilience attenuates the flood–depression relationship, complemented by a machine learning predictive validity assessment (Section 2.8).

### 2.5 Primary Bayesian Multilevel Model

The primary model was a Bayesian multilevel logistic regression fitted using the R package *brms* (version 2.18) ((Bürkner, 2017, 2018)), with *Stan* as the computational backend ((Carpenter et al., 2017)). The model takes the following form:

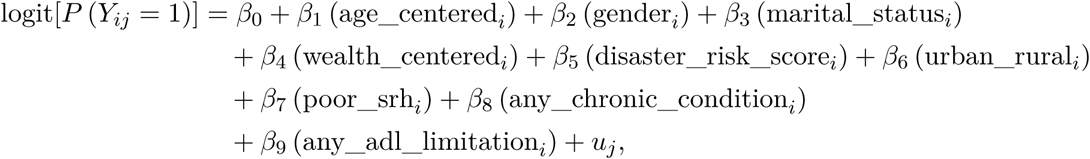

where *Y_ij_*= 1 denotes high depression (CES-D-10 sum ≥ 10) for individual *i* in community *j*, and *u_j_*is a community-level random intercept capturing unobserved between-community heterogeneity.

Age and wealth were mean-centred and scaled by their standard deviations. Binary and categorical predictors were entered using dummy-coded contrasts.

Weakly informative priors were specified: *β*_0_ ∼ N (0, 2.5), *β_k_*∼ N (0, 1) (*k* = 1, . . ., 9), and *σ_u_* ∼ Exponential(1), placing most probability mass on plausible effect sizes for standardised predictors. Four Hamiltonian Monte Carlo chains were run for 2,000 iterations each (1,000 warm-up; adapt_delta = 0.95), with convergence assessed via *R*^^^ *<* 1.01 and effective sample size diagnostics. Posterior summaries are reported as means with 95% credible intervals.

Between-community variance (*σ_u_*^2^) and the intraclass correlation coefficient (ICC) were estimated from the posterior distribution. The ICC on the latent logit scale was computed as:

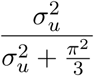

where 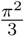 is the level-1 logistic variance under the threshold model interpretation of multilevel logistic regression ((Gelman and Hill, 2006)). Posterior medians and 95% credible intervals are reported for both 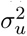 and the ICC.

### 2.6 Variance Decomposition Models

To quantify the extent to which community-level variance in depression risk is attributable to compositional characteristics, disaster exposure, and structural resilience, we fitted a nested sequence of four Bayesian multilevel logistic regression models (M0–M3). M0 is the empty model containing only a random intercept; M1 adds individual covariates; M2 adds the disaster risk score; and M3 adds the structural resilience index. All models share the same random-intercepts specification, prior distributions, and sampling parameters as the primary model (Section 2.5). The proportional change in variance (PCV) at each step was computed relative to M0 to quantify the proportion of between-community variance explained by successive model components. Formal model specifications and the PCV formula are provided in Appendix D.1.

### 2.7 Resilience Moderation Models

Two exposure variables serve distinct roles. The disaster risk score functions as a community-level contextual control capturing chronic ambient hazard (Sections 2.5–2.6); household-level flood exposure is the primary treatment variable in the moderation models, representing the discrete event whose mental health consequences resilience is theorised to buffer.

To test whether community resilience moderates the flood–depression association, two Bayesian multilevel moderation models were fitted: one for structural resilience and one for LCPC. Following the between-within decomposition recommended for clustered data (Gelman and Hill, 2006), flood exposure was decomposed into a between-community component (*x̂_j_*, the community-level proportion of flood-exposed households) and a within-community component (*x_ij_* − *x̂_j_*). The interaction of primary interest is between *x̂_j_* and the resilience index, testing whether communities with higher resilience exhibit a weaker association between aggregate flood prevalence and individual depression risk. The formal model specification is provided in Appendix D.2. Both moderation models used the same prior distributions and sampling settings as the primary model. Conditional effects were evaluated at resilience values of −1, 0, and +1 SD from the mean.

As a robustness check, partially linear Double Machine Learning (DML) models (Bach et al., 2024; Chernozhukov et al., 2018) were estimated with Random Forest nuisance learners and 5-fold cross-fitting, controlling for individual covariates and province; stratum-specific estimates were computed across tertiles of each resilience index.

### 2.8 Machine Learning Predictive Models

To assess the incremental predictive validity of the two resilience indices beyond standard sociodemographic and health predictors, we trained four model variants using both Random Forest and gradient-boosted trees (XGBoost). The baseline feature set comprised demographic characteristics (age, gender, marital status, education level), economic indicators (wealth score, log consumption, disaster risk score), location (urban/rural), and health-related variables (self-rated health, chronic conditions, ADL limitations, and related indicators). Hybrid models extended this baseline by incorporating one of both resilience indices.

To address the circularity arising from deriving LCPC from the same data used in model training (Pagan, 1984; Murphy and Topel, 1985), hybrid models incorporating resilience_re used a province-blocked grouped cross-validation procedure `resilience_re_gkf`). This ensures that no community’s depression outcomes contributed to its own resilience score in the predictive models. The full `resilience_re` derived from Bayesian random intercepts is retained for the moderation and variance decomposition analyses. Full implementation details are provided in the Appendices (Section C.3).

Data were partitioned into stratified training (70%) and test (30%) sets preserving depression prevalence; given 23.6% prevalence, the minority class was upsampled in the training data only, leaving the test set unaltered for unbiased evaluation. Random Forest and XGBoost were implemented with standard settings (Breiman, 2001; Chen and Guestrin, 2016), with categorical variables one-hot encoded and features aligned across partitions. Performance was evaluated primarily by ROC-AUC, with PR-AUC, precision, recall, and F1 computed at a fixed 0.5 threshold.

Geographic generalisability was assessed via leave-one-province-out cross-validation across the 13 IFLS provinces, training on 12 provinces and evaluating on the held-out province with a reduced baseline feature set. With only 13 provinces, spatial buffering for geographic autocorrelation was not feasible; results should be interpreted as a conservative assessment of generalisability.

### 2.9 Sensitivity and Validation Analysis

Two sensitivity analyses were conducted: threshold sensitivity of the binary depression classification across alternative Rasch *θ* cut points, and comparison of the theory-weighted structural resilience index against an equal-weight alternative (*r* = 0.982 across the 312 communities), confirming that substantive conclusions are insensitive to the weighting scheme. Implementation details are provided in the Appendices (Sections C.1 and C.2).

The generated regressor concern arising from deriving LCPC from the same data used in model training is addressed in two ways. For the machine learning predictive models, a grouped *k*-fold cross-validation procedure (`resilience_re_gkf`) ensures that each community’s resilience score is derived from a model never trained on that community’s outcomes, as described in Section 2.8. For the Bayesian moderation analysis, the full random-intercept-based construct is retained; full propagation of uncertainty was not implemented, and those estimates should therefore be interpreted as indicative. Further details are provided in the Appendices (Section C.3).

## 3 Results

### 3.1 Analytical Sample and Descriptive Characteristics

After restricting to individuals aged 15 years with valid CES-D-10 responses residing in original IFLS enumeration areas (EA), the analytical sample comprised 20,303 individuals nested within 312 communities. Restriction to original EAs (defined by the 1993 sampling frame) ensures valid multilevel inference, as these represent the primary probability sampling units.

Depression prevalence (CES-D-10 ≥ 10) was 23.6% (n = 4,785). The sample was 53.7% female, with a mean age of 39.2 years (SD = 15.9). Most respondents were married or cohabiting (70.7%), and 39.4% had primary education or below. Urban residence comprised 28.4% of the sample, reflecting the rural weighting of the original IFLS design. Flood exposure was reported by 10.0% of households (n = 2,021), with community sizes ranging from 1 to 174 individuals (median = 62; IQR: 38–88).

Clear differences were observed between individuals with and without high depression (Table 1). Those with high depression were younger (36.2 vs 40.1 years), more likely to be female (55.9% vs 53.1%), and less likely to be married or cohabiting (64.0% vs 72.7%). They also exhibited higher disaster risk exposure (mean z-score 0.12 vs 0.02) and a greater prevalence of poor self-rated health (31.9% vs 19.0%), chronic conditions (39.0% vs 32.1%), and ADL limitations (21.0% vs 14.3%). Differences in wealth and flood exposure were comparatively modest.

**Table 1:**
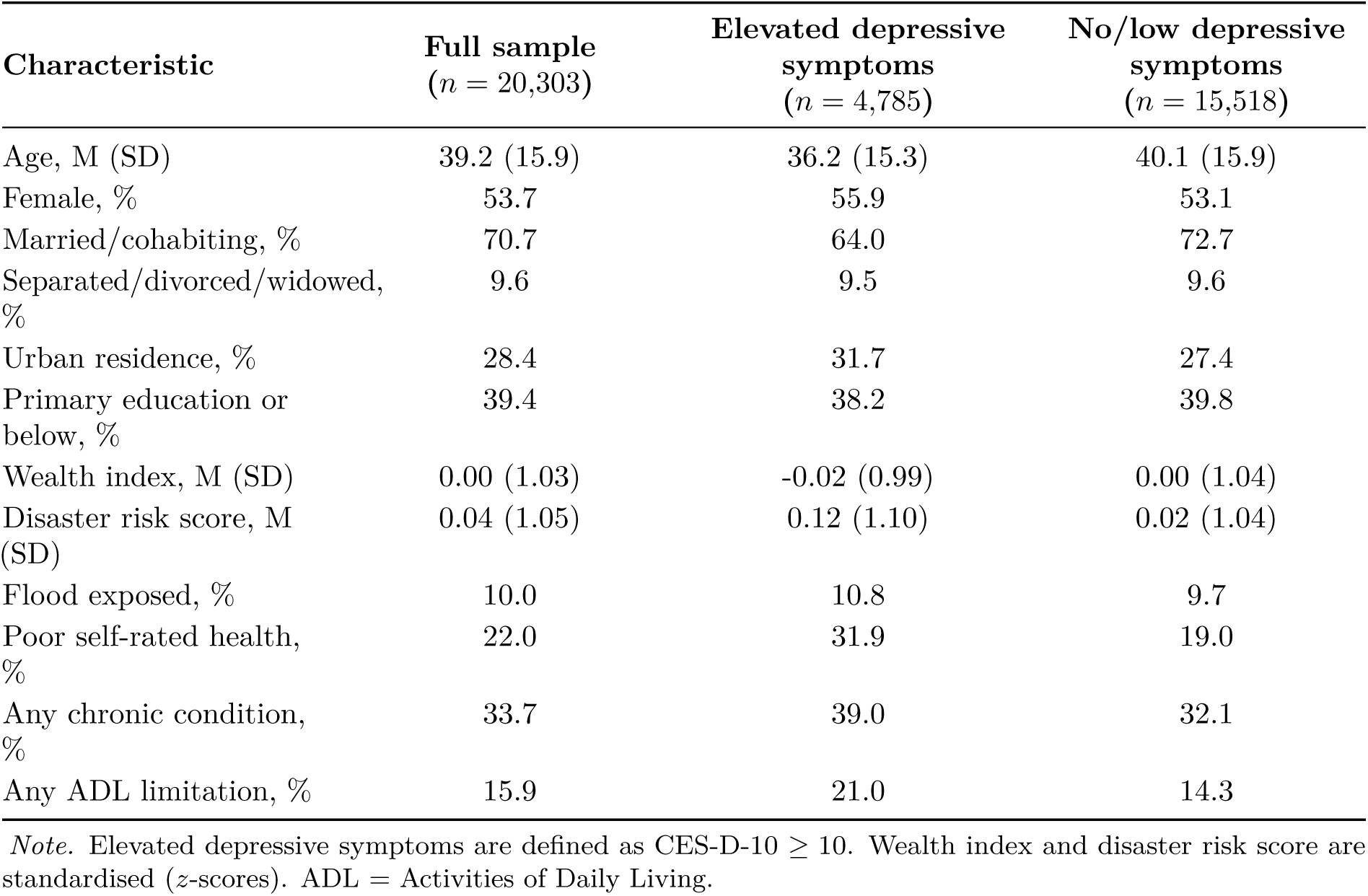
Descriptive statistics of the analytic sample by depression status (*n* = 20,303).

### 3.2 Rasch Partial Credit Model Calibration

The Partial Credit Model calibration yielded person-separation reliability that was moderate (EAP = 0.65; WLE = 0.48), consistent with the brevity of the CES-D-10 and its intended use in community samples (Mair et al., 2025). The *θ* distribution was approximately bell-shaped with slight right skew, reflecting the predominantly non-clinical sample. Sensitivity analyses across alternative percentile-based cutpoints (60th–85th) are reported in Section 3.8.2.

### 3.3 Primary Bayesian Multilevel Model

#### 3.3.1 Convergence and Model Fit

The primary Bayesian multilevel logistic regression model converged satisfactorily across all four Hamiltonian Monte Carlo chains (4,000 post-warm-up draws). All parameters satisfied standard convergence diagnostic (*R*^^^ *<* 1.01), with adequate effective sample sizes. Leave-one-out cross-validation (LOO-CV) indicated stable model performance, with an expected log predictive density (ELPD_LOO_) of −10,363.4 (SE = 74.5), and all Pareto-*k* values below 0.7, suggesting no influential observations.

#### 3.3.2 Individual-Level Predictors

Posterior estimates for fixed effects are presented in Table 2. Health-related variables were the strongest predictors: poor self-rated health approximately doubled the odds of meeting the depression threshold (OR = 2.10, 95% CrI [1.94, 2.28]), with ADL limitation (OR = 1.43) and chronic condition (OR = 1.39) also reliably associated with higher risk, consistent with physical–mental comorbidity documented in LMICs (Patel et al., 2018). Higher wealth (OR = 0.93), greater age (OR = 0.71 per 10 years), and being married or cohabiting (OR = 0.78) were protective. Urban residence was associated with modestly elevated odds (OR = 1.24 [1.02, 1.52]), and sex showed no meaningful association (OR = 1.00), consistent with CES-D-10 studies in South-East Asian samples reporting attenuated sex differences after adjustment. The composite disaster risk score was positively associated with depression risk (OR = 1.06 [1.03, 1.10]), evidence that greater hazard exposure accompanies elevated psychological distress, as examined further in the moderation analyses.

**Table 2:** Posterior estimates from the primary Bayesian multilevel logistic regression model (*n* = 20,273).

| Predictor | Estimate<br>(log-odds) | SD | 95% CrI | OR | 95% CrI |
| --- | --- | --- | --- | --- | --- |
| Intercept | -1.54 | 0.09 | [-1.72, -1.36] | 0.22 | [0.18, 0.26] |
| Age (per 10 years, centred) | -0.34 | 0.02 | [-0.39, -0.30] | 0.71 | [0.68, 0.74] |
| Sex (female vs. male) | 0.00 | 0.04 | [-0.07, 0.07] | 1.00 | [0.93, 1.08] |
| Marital: married/cohabiting | -0.25 | 0.05 | [-0.34, -0.16] | 0.78 | [0.71, 0.85] |
| Marital: separated/divorced/widowed | -0.01 | 0.08 | [-0.17, 0.15] | 0.99 | [0.84, 1.16] |
| Wealth (standardised) | -0.07 | 0.02 | [-0.10, -0.03] | 0.93 | [0.90, 0.97] |
| Disaster risk score | 0.06 | 0.02 | [0.03, 0.10] | 1.06 | [1.03, 1.10] |
| Urban (vs. rural) | 0.22 | 0.10 | [0.02, 0.42] | 1.24 | [1.02, 1.52] |
| Urban (unknown vs. rural) | 0.00 | 0.09 | [-0.18, 0.19] | 1.00 | [0.84, 1.21] |
| Poor self-rated health | 0.74 | 0.04 | [0.66, 0.82] | 2.10 | [1.94, 2.28] |
| Any chronic condition | 0.33 | 0.04 | [0.25, 0.40] | 1.39 | [1.28, 1.49] |
| Any ADL limitation | 0.36 | 0.05 | [0.27, 0.45] | 1.43 | [1.31, 1.56] |

*Note.* Estimate = posterior mean on the log-odds scale; SD = posterior standard deviation; CrI = credible interval; OR = odds ratio. Reference categories are male, unmarried, and rural. Age is centred at the sample mean (39.2 years), and wealth is standardised ( $M = 0$ , $SD = 1$ ). The community-level random-intercept SD was $\sigma_u = 0.443$ (95% CrI [0.389, 0.501]). All $\hat{R} < 1.01$ .
ICC = 0.061). Individual covariates (M1) reduced the variance by 8.1%, the disaster risk score (M2) added only a marginal further reduction (PCV = 9.5%), and structural resilience (M3) reduced the variance to $\sigma_u^2 = 0.186$ (PCV = 13.2%), an independent contribution to explaining community differences, though the majority of variance remains unexplained, pointing to residual contextual factors not captured by observed covariates.

#### 3.3.3 Community-Level Variance

The estimated community-level random intercept standard deviation was *σ_u_*= 0.443 (95% CrI [0.389, 0.501]), corresponding to *σ_u_*^2^ = 0.196 and an ICC of 0.056 on the latent logistic scale: approximately 6% of total variance in depression risk is attributable to between-community differences, supporting the use of the multilevel framework.

### 3.4 Community Variance Decomposition (M0–M3)

A sequence of nested Bayesian multilevel models quantified how far between-community variation is explained by individual composition, disaster exposure, and structural resilience (Table 3); PCV is computed relative to the empty model. M0 confirmed non-negligible clustering (*σ*^2^ = 0.215;

**Table 3:** Community-level variance decomposition across nested Bayesian multilevel models (M0–M3).

| Model | Specification | $\sigma_u^2$ | ICC | PCV (%) | Interpretation |
| --- | --- | --- | --- | --- | --- |
| **M0** | Empty (null) | 0.215 | 0.061 | — | Unconditional community heterogeneity |
| **M1** | + Individual covariates | 0.197 | 0.057 | 8.1 | Individual composition explains approximately 8% of community variance |
| **M2** | + Disaster risk score | 0.194 | 0.056 | 9.5 | Minimal additional variance explained by disaster risk |
| **M3** | + Structural resilience | 0.186 | 0.054 | 13.2 | Structural resilience explains further community variance |

ICC = 0.061). Individual covariates (M1) reduced the variance by 8.1%, the disaster risk score (M2) added only a marginal further reduction (PCV = 9.5%), and structural resilience (M3) reduced the variance to *σ*^2^ = 0.186 (PCV = 13.2%), an independent contribution to explaining community differences, though the majority of variance remains unexplained, pointing to residual contextual factors not captured by observed covariates.

### 3.5 Community Resilience: Descriptive Patterns

#### 3.5.1 Structural Resilience

The structural resilience index varied across the 312 IFLS original communities (range: −1.83 to 3.43; mean = −0.18, SD = 0.67). Convergent validity was supported by principal component analysis: the first principal component explained 53.5% of total variance and correlated at *r* = 0.994 with the composite index, with loadings highest for wealth and consumption; loading patterns and scree plots are shown in the Supplementary Materials (Section C.2). Depression prevalence declined only modestly across resilience quartiles (25.0% in Q1 to 22.2% in Q4), with substantial within-quartile heterogeneity (Figure 1); several high-resilience communities exhibit elevated depression rates, underscoring the distinction between structural capacity and realised resilience and motivating the LCPC measure examined below.

**Figure 1:**
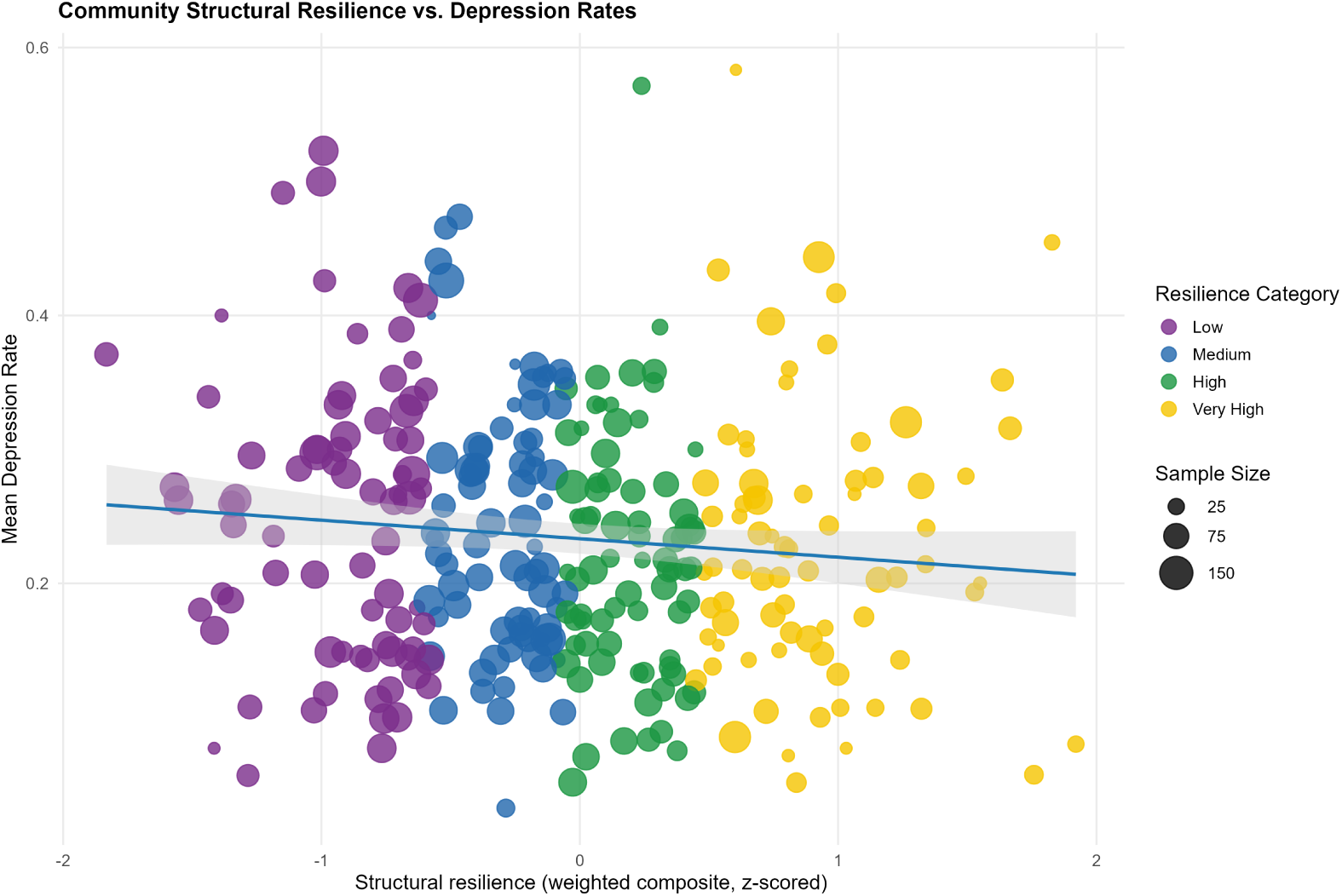
Community structural resilience versus mean depression rate across 312 IFLS original communities. Point size is proportional to community sample size; colour indicates resilience quartile. Source: Author’s analysis of IFLS-5 data.

#### 3.5.2 Latent Community Protective Capacity

LCPC scores (`resilience_re` = scale(-*u_j_*)) ranged from −3.35 to 2.37 across 312 communities (*Mean* ≈ 0.00, SD = 1.10), reflecting the inverted and standardised community random intercepts from the primary Bayesian model. Higher values indicate communities whose observed depression rates were lower than predicted by individual-level covariates alone and can be interpreted as a proxy for latent protective contextual capacity not captured by the structural resilience index.

The correlation between structural resilience and Latent Community Protective Capacity at the community level was *r* = 0.155, confirming that the two indices are weakly correlated and empirically distinct and capture distinct dimensions of community protective capacity. This low correlation is theoretically important: structurally well-resourced communities are not necessarily those exhibiting lower-than-expected depression once individual-level composition is accounted for.

In contrast to the weak and diffuse relationship observed for structural resilience (Figure 1), LCPC exhibits a strong and near-linear negative association with depression prevalence (Figure 2). Depression prevalence declines sharply across LCPC quartiles, from 35.6% in Q1 to 12.4% in Q4. The relationship is characterised by tight clustering around the fitted regression line and clear separation between quartiles, indicating substantially greater alignment with observed community-level variation in depression.

**Figure 2:**
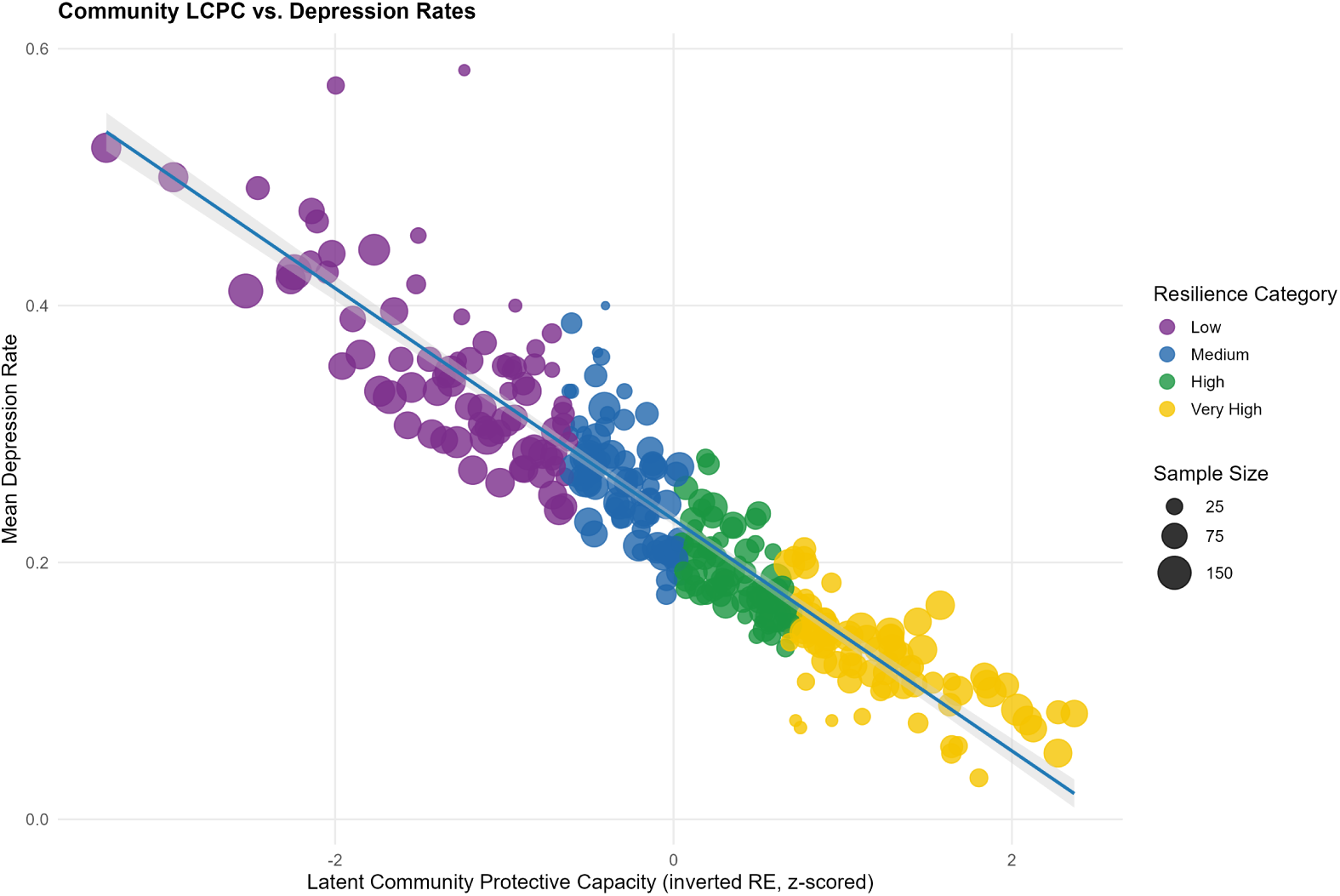
LCPC versus mean depression rate across 312 IFLS original communities. Point size is proportional to community sample size; colour indicates resilience quartile.

Because LCPC is constructed from community-level random effects, part of its association with the outcome is mechanical; it is precisely this property that allows it to quantify unexplained contextual variation, approximating realised protective capacity in contrast to the potential capacity represented by structural resilience. The stark contrast between Figures 1 and 2 highlights the importance of distinguishing these two dimensions when assessing resilience in population mental health.

### 3.6 Resilience Moderation of Flood–Depression Association

#### 3.6.1 Structural Resilience Moderation

The structural moderation model estimated the interaction between community flood prevalence (*x̂_j_*) and structural resilience (`resilience_struct`) as a predictor of individual depression probability. The posterior mean for the interaction term (*x̂_j_* × `resilience_struct`) was 0.041 (SE = 0.050, 95% CrI [−0.057, 0.140]), providing no evidence of a reliable moderation effect. The credible interval spans zero and remains wide, indicating substantial posterior uncertainty and precluding meaningful directional interpretation.

Conditional effects evaluated at −1 SD, the mean, and +1 SD of structural resilience show near-parallel predicted probability curves with extensively overlapping 95% credible intervals (Figure 3), indicating that structural resilience, as operationalised here, does not meaningfully alter the flood–depression association; consistent with its interpretation as baseline resource capacity rather than a realised protective mechanism.

**Figure 3:**
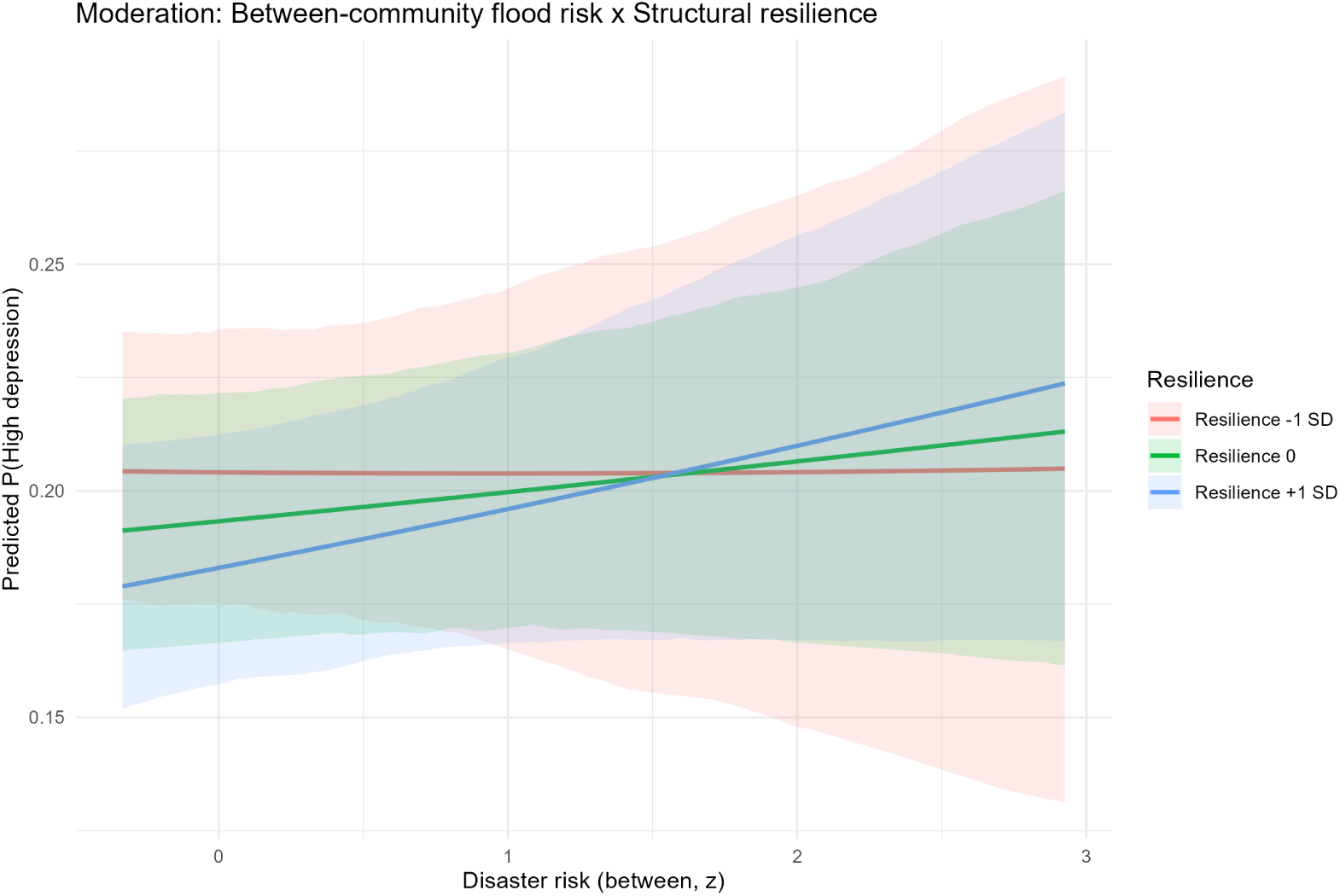
Conditional effects of between-community flood prevalence on predicted depression probability at three levels of structural resilience (−1 SD, mean, +1 SD). Shaded bands represent 95% posterior credible intervals.

#### 3.6.2 LCPC Moderation

The LCPC moderation model substituted `resilience_re for resilience_struct` while maintaining an otherwise identical specification. The posterior mean for the interaction term (*x̂_j_* × `resilience_re`) was −0.023 (SE = 0.036, 95% CrI [-0.093, 0.047]). The credible interval spans zero and the point estimate is small in absolute magnitude (OR scale: exp (−0.023) ≈ 0.977 per unit increase in community flood prevalence at mean LCPC). Nonetheless, the direction of the interaction is consistent with the buffering hypothesis: the negative sign indicates that communities with higher LCPC show a weaker positive association between community-level flood risk and individual depression probability. Approximately 74% of posterior mass falls below zero, indicating a directional tendency consistent with the moderation hypothesis, albeit with substantial uncertainty.

The contrast between the two moderation results is itself informative: the structural interaction is positive and directionally inconsistent with buffering (0.041, CrI [−0.057, 0.140]), while the LCPC interaction is negative and aligned with theoretical expectation (−0.023, CrI [−0.093, 0.047]). The clear vertical separation of predicted probability curves in Figure 4 indicates a strong main effect of LCPC on baseline depression risk, with comparatively modest differences in slope: strong baseline effects but limited effect modification.

**Figure 4:**
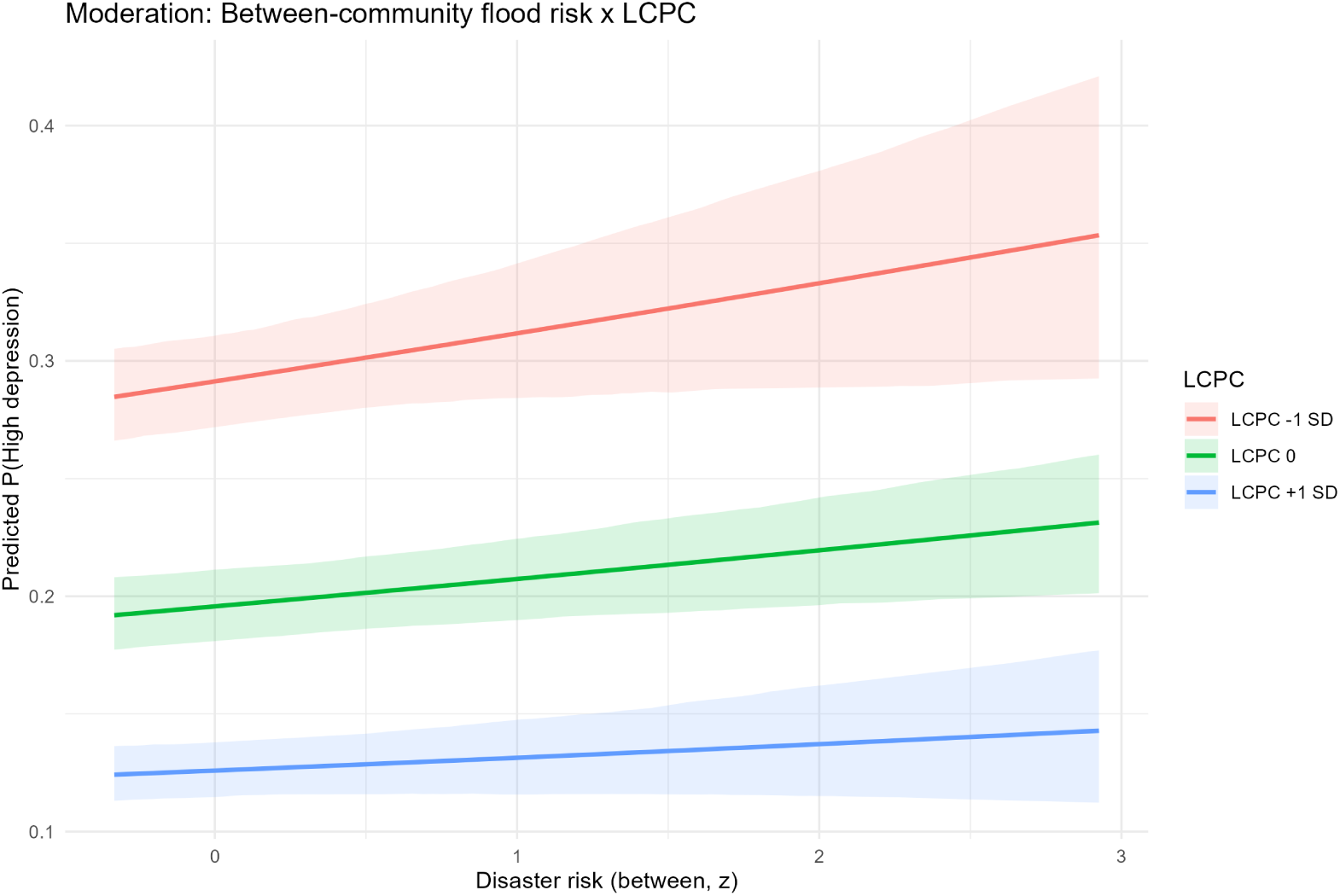
Conditional effects of between-community flood prevalence on predicted depression probability at three levels of LCPC (−1 SD, mean, +1 SD). Shaded bands represent 95% posterior credible intervals. Source: Author’s analysis of IFLS-5 data.

#### 3.6.3 Double Machine Learning Robustness Check

Partially linear DML models with Random Forest nuisance learners and 5-fold cross-fitting provide a robustness check on the moderation findings under weaker parametric assumptions than the Bayesian models (Chernozhukov et al., 2018).

For the structural resilience DML model, the estimated average treatment effect of flood exposure on depression was *θ* = 0.021 (SE = 0.015, *t* = 1.41, *p* = 0.160), indicating a positive but imprecisely estimated association between flood exposure and depression probability after partialling out covariates. Stratum-specific estimates were consistent across low, mid, and high structural resilience communities (*θ* = 0.000, 0.025, 0.015 respectively), with all standard errors overlapping zero, providing no strong evidence that the effect of disaster risk on depression differed systematically by structural resilience level (Figure 5).

**Figure 5:**
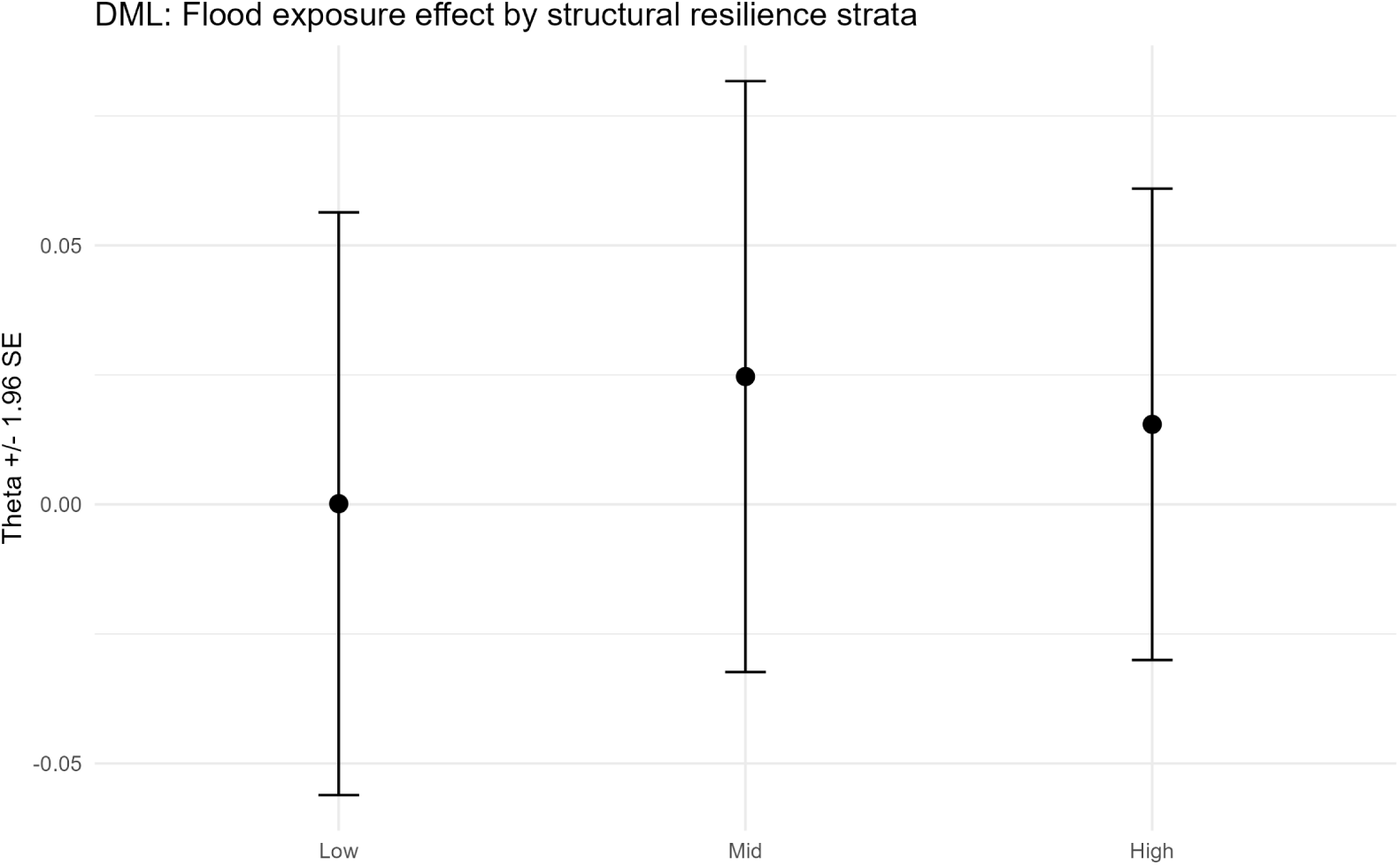
DML stratum-specific estimates of the flood exposure effect on depression probability (*θ* ± 1.96 SE) by structural resilience tertile. Point estimates are near-zero across low, mid, and high strata with overlapping confidence intervals, providing no evidence of systematic heterogeneity in the flood–depression association by structural resilience level. Source: Author’s analysis of IFLS-5 data.

For the LCPC DML model, the overall treatment effect was similarly small (*θ* = 0.019, SE = 0.015, *t* = 1.28, *p* = 0.199). Stratum-specific estimates suggested a directionally consistent pattern: communities in the low LCPC stratum exhibited the largest positive treatment effect (*θ* = 0.053, SE = 0.033), while mid- and high-resilience communities showed near-zero effects (*θ* = 0.003 and *θ* = −0.002 respectively). Although none of these stratum-level contrasts achieves conventional significance thresholds, the monotonic decline in the treatment effect across resilience levels (Figure 6) is consistent with a directional tendency that LCPC buffers the flood–depression pathway.

**Figure 6:**
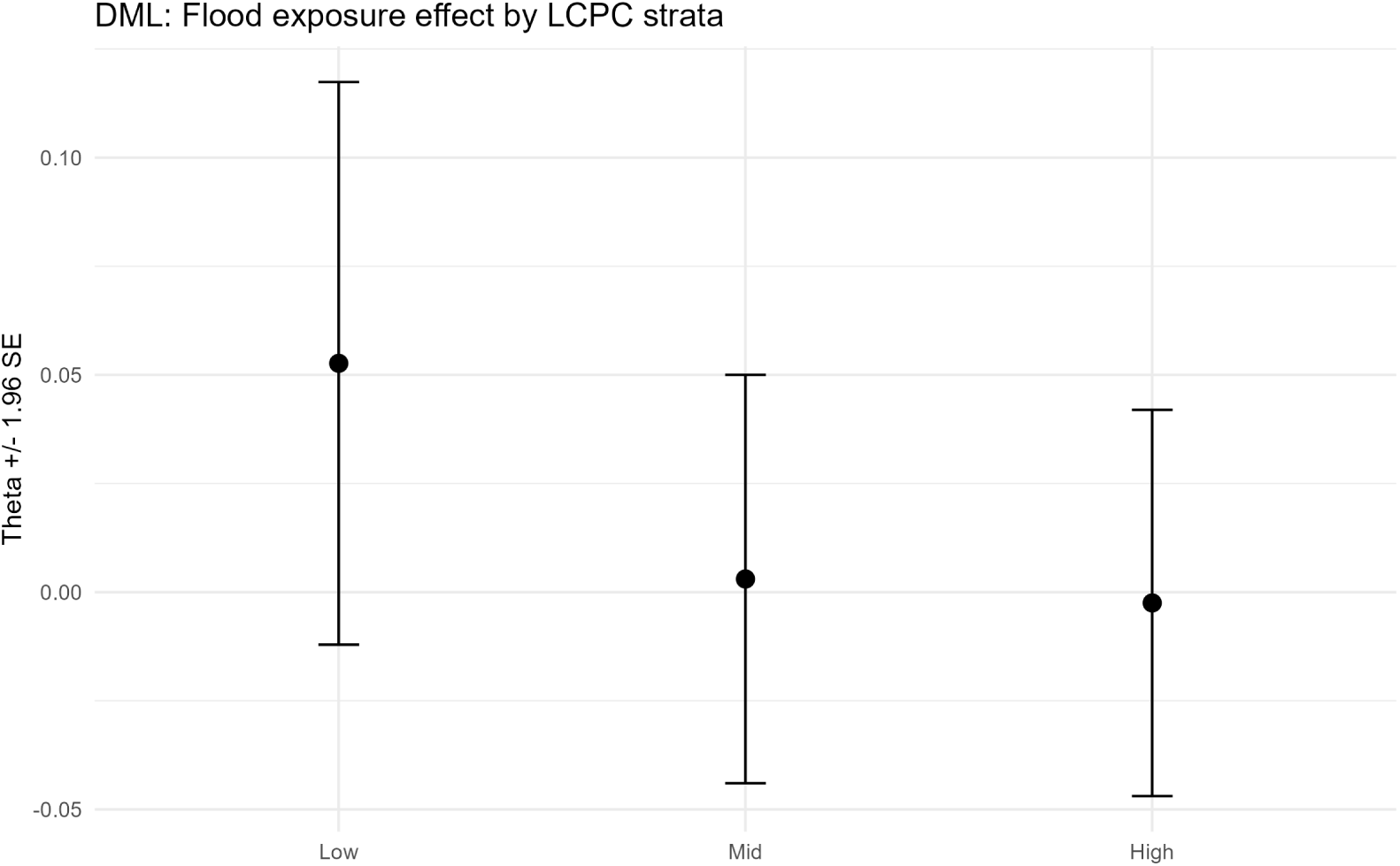
DML stratum-specific estimates of the flood exposure effect on depression probability (*θ* ± 1.96 SE) by LCPC tertile. The point estimate declines monotonically from the low to the high LCPC stratum (*θ* = 0.053, 0.003, and −0.002 respectively), consistent with the hypothesis that communities with greater latent protective capacity exhibit a weaker flood–depression association. Source: Author’s analysis of IFLS-5 data.

Taken together, the DML results reinforce the earlier findings: structural resilience shows no meaningful effect modification, whereas LCPC exhibits a weak but coherent pattern aligned with the hypothesised buffering mechanism.

Across modelling approaches, a consistent pattern emerges: structural resilience shows no meaningful effect modification, whereas LCPC exhibits a weak but coherent buffering signal, a negative interaction with approximately 74% of posterior mass below zero and monotonically declining DML stratum estimates, alongside a strong main effect on baseline depression risk. Latent contextual capacity is thus more closely aligned with both baseline mental health outcomes and their sensitivity to environmental shocks than structural resource composites. This distinction is examined further through predictive modelling below.

### 3.7 Machine Learning Predictive Validity

#### 3.7.1 Classification Performance

These analyses evaluate whether the distinction between weak moderation and strong baseline effects of resilience is reflected in out-of-sample predictive performance.

Table 4 presents ROC-AUC, PR-AUC, precision, recall, and F1 for all eight model variants (four feature sets × two algorithms) evaluated on the 30% held-out test set. Across all models, ROC-AUC ranged from 0.635 (RF Baseline) to 0.708 (XGBoost Hybrid-RE), indicating moderate discriminative ability consistent with the complexity of community-level mental health prediction from survey covariates.

**Table 4:**
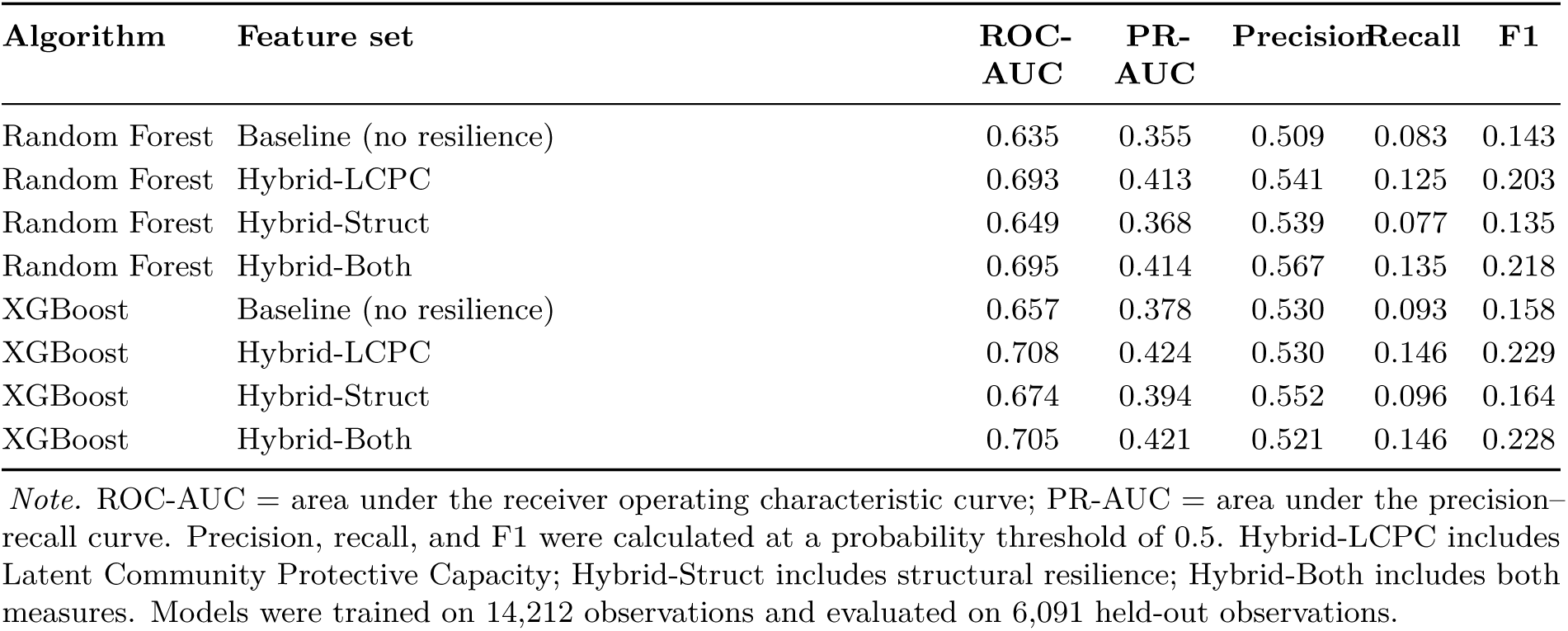
Machine-learning classification performance by algorithm and feature set in the held-out test sample.

| Algorithm | Feature set | ROC-AUC | PR-AUC | Precision | Recall | F1 |
| --- | --- | --- | --- | --- | --- | --- |
| Random Forest | Baseline (no resilience) | 0.635 | 0.355 | 0.509 | 0.083 | 0.143 |
| Random Forest | Hybrid-LCPC | 0.693 | 0.413 | 0.541 | 0.125 | 0.203 |
| Random Forest | Hybrid-Struct | 0.649 | 0.368 | 0.539 | 0.077 | 0.135 |
| Random Forest | Hybrid-Both | 0.695 | 0.414 | 0.567 | 0.135 | 0.218 |
| XGBoost | Baseline (no resilience) | 0.657 | 0.378 | 0.530 | 0.093 | 0.158 |
| XGBoost | Hybrid-LCPC | 0.708 | 0.424 | 0.530 | 0.146 | 0.229 |
| XGBoost | Hybrid-Struct | 0.674 | 0.394 | 0.552 | 0.096 | 0.164 |
| XGBoost | Hybrid-Both | 0.705 | 0.421 | 0.521 | 0.146 | 0.228 |

*Note.* ROC-AUC = area under the receiver operating characteristic curve; PR-AUC = area under the precision-recall curve. Precision, recall, and F1 were calculated at a probability threshold of 0.5. Hybrid-LCPC includes Latent Community Protective Capacity; Hybrid-Struct includes structural resilience; Hybrid-Both includes both measures. Models were trained on 14,212 observations and evaluated on 6,091 held-out observations.

PR-AUC values (0.355–0.423) were substantially lower than ROC-AUC, as expected at 23.6% class prevalence (Davis and Goadrich, 2006); recall at the fixed 0.5 threshold is correspondingly low, reflecting a precision-oriented operating point rather than deployment-optimised classification.

The addition of resilience variables produced consistent improvements in both metrics: including resilience_re alone improved XGBoost ROC-AUC by 5.1 percentage points (0.657 → 0.708) and PR-AUC by 4.5 points (0.378 → 0.423) over the no-resilience baseline. Structural resilience produced smaller but consistent gains (ROC-AUC +1.7pp, PR-AUC +1.6pp), while combining both indices yielded performance nearly identical to LCPC alone. This pattern indicates that resilience_re captures the dominant community-level signal relevant for prediction, with limited additional marginal contribution from the structural index.

The objective of these models is not maximal accuracy but incremental predictive contribution: overall discrimination (ROC-AUC ≈ 0.70) is consistent with prior survey-based population mental health prediction, and the key result is the systematic improvement associated with resilience variables, confirming that community-level resilience captures information about depression risk not already encoded in individual characteristics, complementing rather than replacing inferential methods (Akindejoye et al., 2025).

#### 3.7.2 Feature Importance and SHAP Interpretability

Feature importance rankings from both algorithms consistently placed resilience_re (the province-blocked GKF approximation; see Section 2.8) among the top predictors in hybrid configurations. To prevent target leakage, the CES-D-10 sum score, Rasch severity estimate, and all item responses were excluded from the feature matrix. Among exogenous predictors, poor self-rated health, age, and resilience_re dominated; the correlation of resilience_re with the outcome (*r* = 0.218) was the highest of any community-level variable.

SHAP (SHapley Additive exPlanations; Lundberg and Lee, 2017) values for the XGBoost Hybrid-Both model confirm resilience_re as the single largest contributor to predictions (Figure 7). At the individual level, above-average community LCPC exerts a substantial downward correction on predicted risk even where health and sociodemographic factors push it upward, and the SHAP dependence relationship for LCPC is monotonically negative, whereas that for structural resilience is diffuse and centred near zero, consistent with LCPC’s dominant share of feature importance and with the broader finding that observed structural resources only partially capture the protective contextual capacity reflected in LCPC (individual-level SHAP waterfall and dependence plots are provided in the Appendices, Section D.3).

**Figure 7:**
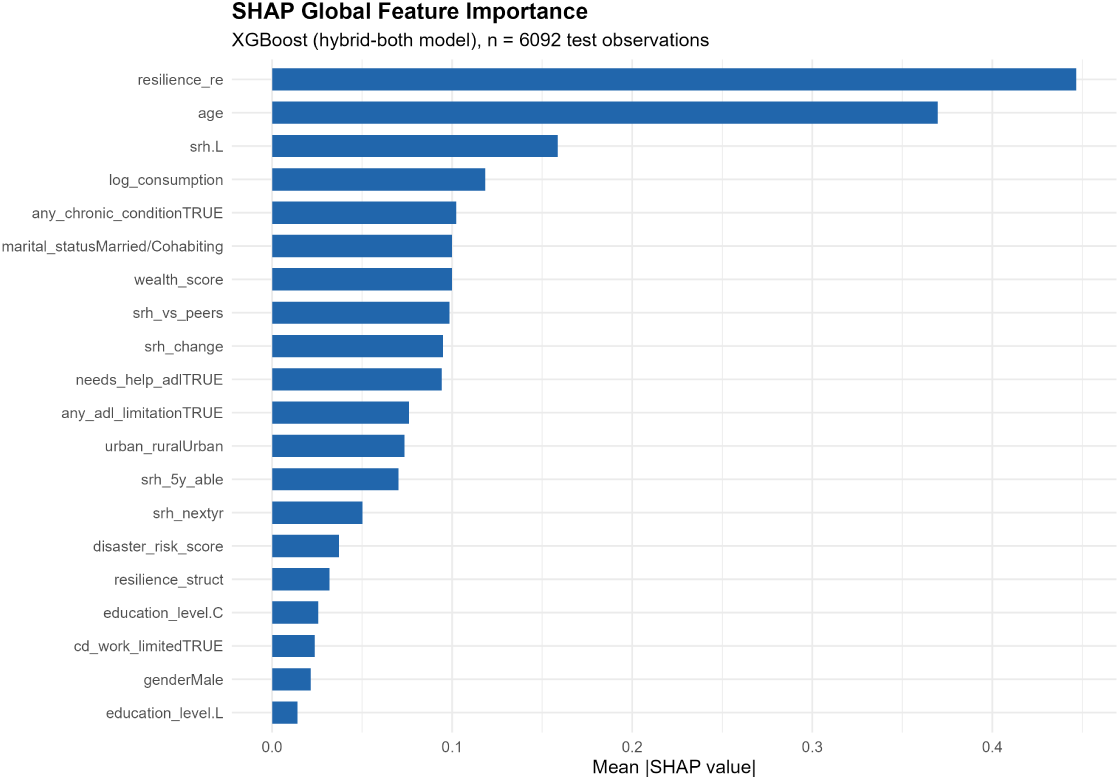
SHAP global feature importance for the XGBoost Hybrid-Both model (n = 6,092 test observations). Bars show mean absolute SHAP value. `resilience_re` (LCPC) is the single highest-importance feature, substantially exceeding all individual-level predictors. Source: Author’s analysis of IFLS-5 data.

### 3.8 Sensitivity and Validation Analyses

#### 3.8.1 Geographic External Validity

Leave-one-province-out cross-validation across all 13 provinces (Table 5) yielded ROC-AUC from 0.577 (West Nusa Tenggara) to 0.668 (Yogyakarta), mean 0.614. The narrow range and absence of provinces approaching chance performance indicate stable, non-trivial predictive signal across geographically and socioeconomically diverse regions, suggesting broadly generalisable patterns rather than location-specific artefacts. Resilience indices were intentionally excluded to provide a conservative test based solely on individual covariates; reported performance therefore represents a lower bound, and including resilience variables would be expected to improve discrimination. Together, these results indicate that while absolute performance is moderate, the contribution and robustness of community-level resilience, particularly resilience_re, are consistently supported.

**Table 5:** Leave-one-province-out cross-validation results (Random Forest, baseline feature set). Models were trained on all remaining provinces and evaluated on the held-out province.

| Code | Province | Test $n$ | Positive | Negative | AUC |
| --- | --- | --- | --- | --- | --- |
| 12 | North Sumatra | 1,343 | 274 | 1,069 | 0.609 |
| 13 | West Sumatra | 990 | 201 | 789 | 0.615 |
| 16 | South Sumatra | 830 | 146 | 684 | 0.629 |
| 18 | Lampung | 968 | 165 | 803 | 0.610 |
| 31 | DKI Jakarta | 1,388 | 315 | 1,073 | 0.630 |
| 32 | West Java | 3,131 | 971 | 2,160 | 0.587 |
| 33 | Central Java | 2,775 | 643 | 2,132 | 0.616 |
| 34 | DI Yogyakarta | 1,228 | 219 | 1,009 | 0.668 |
| 35 | East Java | 2,948 | 458 | 2,490 | 0.637 |
| 51 | Bali | 1,111 | 314 | 797 | 0.588 |
| 52 | West Nusa Tenggara | 1,712 | 535 | 1,177 | 0.577 |
| 63 | South Kalimantan | 850 | 210 | 640 | 0.594 |
| 73 | South Sulawesi | 1,029 | 334 | 695 | 0.615 |
| **Mean (across provinces)** |  | **1,485** | **368** | **1,116** | **0.614** |
| **Range** |  | 830–3,131 | 146–971 | 640–2,490 | 0.577–0.668 |

*Note.* Province codes follow the Indonesian Badan Pusat Statistik (BPS) two-digit classification. Positive = number of elevated-depressive-symptom cases (CES-D-10 $\geq 10$ ) in the test fold; Negative = number of non-cases. AUC = area under the receiver operating characteristic curve. The baseline feature set comprised age, gender, wealth score, disaster risk score, poor self-rated health, any chronic condition, and any ADL limitation.

#### 3.8.2 Measurement Sensitivity Analyses

Threshold sensitivity analyses across six alternative CES-D *θ* percentile cutpoints (60th–85th) confirmed stable performance across reasonable threshold choices (Supplementary Materials, Section S3.1). The primary theory-weighted structural resilience index correlated at *r* = 0.982 with an equal-weight alternative, confirming that substantive conclusions are insensitive to the specific weighting scheme (Appendices Section C.1).

## 4 Discussion

### 4.1 Principal Findings

This study examined community-level determinants of depression risk following flood exposure among 20,303 Indonesian adults nested within 312 original IFLS communities. Three interconnected findings emerge. First, community context accounts for a meaningful, if modest, share of the variance in depression risk (ICC = 6.1%), independent of individual-level predictors. Second, structural resilience, a theoretically weighted composite of community wealth, consumption, education, urbanisation, and social trust, accounts for an additional 3.7 percentage points of between-community variance beyond flood exposure (PCV M2→M3), indicating that material and social resources genuinely stratify community mental health outcomes. Third, neither resilience index moderates the flood–depression relationship with posterior certainty: the structural interaction is effectively null (*interaction coefficient* = 0.041, 95% CrI [-0.057, 0.140]), while the LCPC moderation is directional but inconclusive (*interaction coefficient* = −0.023, 95% CrI [-0.093, 0.047]), with 74% of posterior mass below zero. Machine learning analyses corroborate the predictive signal of resilience variables, which rank among the highest-importance features in the best-performing XGBoost model (ROC-AUC = 0.708).

These findings indicate that resilience operates primarily through baseline differences in mental health risk across communities rather than through strong effect modification of flood exposure, an interpretation consistent with the DML and predictive analyses.

### 4.2 Interpreting the Findings

The ICC of 6.1% confirms that community membership is a non-trivial determinant of depression risk over and above individual characteristics (Kawachi and Berkman, 2001; Norris et al., 2008). Crucially, the near-independence of structural resilience and LCPC (*r* = 0.155) is a central empirical finding: structurally well-resourced communities are not automatically those exhibiting lower-than-expected depression given their composition, implying that the processes most relevant to psychological outcomes are not fully captured by observable socioeconomic indicators.

The structural index was validated against an equal-weights variant and the first principal component of its constituents (*r* = 0.994), confirming the theoretical weights do not distort the latent structure; its alignment with multi-capital framings (Ngai et al., 2022) lends cross-contextual credibility, while the positive urban–depression association (OR = 1.24) reinforces that urbanisation is not straightforwardly protective in Indonesia (Hutter and Bailey, 2022).

The null structural moderation result is unlikely to be fully explained by measurement imprecision or insufficient power: the credible interval is relatively narrow and DML robustness checks confirm a near-zero average treatment effect of flood exposure on depression (*θ* = 0.021, p = 0.160). Three substantive explanations are plausible. First, the cross-sectional design captures chronic flood hazard rather than acute event timing, precluding detection of longitudinal buffering that may only emerge under temporally resolved exposure and recovery dynamics. Second, flood exposure is concentrated among economically marginalised communities, which also score lowest on structural resilience, leaving insufficient variation in the joint distribution to identify an interaction. Third, material resources may buffer economic but not psychological recovery, as documented elsewhere in the disaster literature (Golitaleb et al. (2022)). The directional LCPC moderation result, 74% of posterior mass below zero, is consistent with the interpretation that unmeasured institutional and social processes, rather than observable material resources, may reflect more proximate determinants of psychological resilience. This interpretation is further supported by the stratified DML analysis, which suggests a declining pattern of flood effects across LCPC strata, but no systematic pattern across structural resilience levels. Hutter and Bailey (2022) make precisely this argument: the indicators most consequential for resilience outcomes are those hardest to quantify and least represented in existing frameworks.

At the individual level, poor self-rated health (OR = 2.10) dominates the fixed-effects model, consistent with the bidirectional relationship between physical and mental health under disaster stress. The protective age gradient (OR = 0.71 per 10-year increment) may reflect that younger adults bear disproportionate economic and caregiving burdens following floods. The predictive analyses provide complementary evidence of the incremental value of resilience measures: while overall discrimination remains moderate (ROC-AUC ≈ 0.70), consistent with prior work using survey-based mental health data, the inclusion of resilience variables yields systematic improvements in both ROC-AUC and PR-AUC. This reinforces that the key contribution lies in the additional explanatory and predictive signal captured by community-level resilience, rather than in absolute predictive accuracy.

## 5 Policy Implications

The central policy-relevant finding of this study is that a large share of the community-level capacity that protects against post-flood depression is invisible to the resource-based indicators on which conventional vulnerability assessment relies. Structural resources: wealth, consumption, education, urbanisation, and social trust, account for only a modest fraction of between-community variation in depression risk, while the greater part is captured by a latent measure (LCPC) derived from the outcomes themselves. For disaster policy, this has a direct consequence: targeting psychosocial resources solely on the basis of observable material and demographic disadvantage will systematically misallocate support, directing it away from communities that are materially better off yet, for reasons of governance, social process, or institutional capacity, absorb the psychological impact of flooding less well than their resources would predict.

### 5.1 Potential users and applications

The measure is most readily actionable for national and sub-national disaster management agencies (in the Indonesian case, the Badan Nasional Penanggulangan Bencana, BNPB, and its provincial counterparts), health ministries responsible for mental health service planning, and humanitarian and non-governmental actors delivering post-disaster psychosocial support. Three uses are plausible. First, *preparedness targeting*: because LCPC can be estimated from routinely collected household survey data, it can be computed in advance of flood events to flag communities where post-flood mental health need is likely to exceed what resource-based indices anticipate, informing the pre-positioning of psychosocial first aid capacity and referral pathways. Second, *resource allocation during response and early recovery*, where LCPC offers a complementary screening layer alongside physical damage and exposure assessments. Third, *monitoring and evaluation*, where changes in community-level protective capacity over successive survey waves could serve as an indicator of whether resilience-building interventions are achieving their intended effect, aligning with the Sendai Framework’s emphasis on measurable progress in psychosocial recovery.

### 5.2 Implementation requirements and barriers

Realising these uses depends on infrastructure and capacity that vary widely across disaster-prone states. The approach requires periodic, geographically identifiable household survey data with mental health and socioeconomic modules, and the analytical capacity to fit multilevel models and generate out-of-sample community scores. Many low- and middle-income countries maintain survey programmes of the necessary scale, but fewer have the standing analytical infrastructure to operationalise them for real-time decision support; investment in that capacity, or partnership with research institutions, would be a precondition. A governance question also follows: because LCPC is a residual, model-derived quantity rather than a directly observed one, its use in resource allocation would require transparent methodology, periodic revalidation, and clear institutional ownership of the measure to avoid opaque or unaccountable targeting.

### 5.3 Policy limitations and wider significance

Two cautions bound these implications. The estimates here are associational: LCPC identifies where elevated need is likely to concentrate, not why, and should inform the *targeting* of interventions rather than substitute for local assessment of their content. And because LCPC is derived residually, a low score signals that a community underperforms relative to its resources without identifying the cause, healthcare access, governance quality, or social cohesion may each be implicated, so it is best treated as a screening signal that directs further inquiry, not as a diagnosis. Validating the measure against independent, real-world post-flood mental health outcomes, and testing its portability to higher-income settings such as the United Kingdom where the underlying survey infrastructure differs, is a necessary next step before the approach is embedded in operational policy.

Although developed on Indonesian data, the contribution is not specific to Indonesia. The analytical logic: recovering latent protective capacity from the survey infrastructure a state already maintains, and using it to correct the blind spots of resource-based vulnerability indices, applies wherever comparable data exist, and is directly relevant to the design of analogous tools in other low- and middle-income settings facing intensifying flood hazard. It also establishes the methodological template for the higher-income, UK-based extension that forms the next stage of this research programme, where the same question: whether observable resources adequately capture community protective capacity, carries equal weight for health-system planning.

## 6 Conclusion

This study asked how community resilience can be quantified in large-scale household survey data, how far structural resources explain between-community heterogeneity in post-flood depression risk, and whether resilience measures improve inferential and predictive identification of mental health vulnerability. Two complementary constructs were developed: a theoretically weighted structural resilience index aggregated from observable community resources, and a Latent Community Protective Capacity (LCPC) index derived from the inverted community random intercepts of the Bayesian multilevel model. Their near-orthogonality (*r* = 0.155) indicates that observable resource composites alone are insufficient to characterise community resilience. Structural resilience accounts for approximately 3.7 percentage points of between-community variance beyond individual covariates and flood exposure (total PCV = 13.2%), yet roughly 87% of the original between-community variance remains unexplained, residual contextual capacity that LCPC is designed to approximate. Neither index moderates the flood–depression association with posterior certainty, though the LCPC result is directionally consistent with buffering; predictively, adding LCPC improves XGBoost ROC-AUC by 5.1 percentage points over the no-resilience baseline and dominates SHAP feature importance.

Several limitations bear on interpretation. The cross-sectional design precludes causal interpretation and temporal dynamics; the CES-D-10 is a screening rather than diagnostic instrument with moderate reliability (EAP ≈ 0.65); the flood exposure measure omits magnitude, duration, and physical loss; and the structural index uses theoretically rather than empirically optimised weights, with the weak urban loading suggesting the BRIC framework may require adaptation for Southeast Asian settings. LCPC is a residual construct: it may capture omitted contextual factors: healthcare access, governance quality, environmental conditions, cultural norms, alongside protective processes, and its generated-regressor character is addressed for the predictive models through province-blocked grouped cross-validation (Section 2.8) but retained without full uncertainty propagation in the Bayesian moderation analysis, whose estimates should therefore be read as indicative. Future work should prioritise longitudinal linkage of IFLS waves to estimate depression trajectories around flood events, qualitative inquiry in communities combining high LCPC with low structural resources, and extension of the framework to other disaster-prone low- and middle-income settings where comparable survey data exist.

Taken together, the findings suggest that the community processes most protective against post-flood depression are likely those hardest to quantify through conventional survey indicators, a conclusion with direct consequences for how disaster policy identifies and supports at-risk populations (Section 5). By recovering this latent protective capacity from survey infrastructure that disaster-prone states already maintain, the approach offers a route to more accurately targeted psychosocial preparedness and response, consistent with the health priorities of the Sendai Framework, while the methodological framework itself transfers to other settings confronting escalating climate-related hazard exposure.

## Ethics statement

This study is a secondary analysis of fully anonymised, publicly available data from the Indonesia Family Life Survey Wave 5 (IFLS-5). The IFLS-5 survey protocols and instruments were reviewed and approved by Institutional Review Boards at the RAND Corporation (United States) and Universitas Gadjah Mada (Indonesia), and informed consent was obtained from all respondents at the time of data collection. No additional ethical approval was required for the present secondary analysis of de-identified public data; all procedures complied with relevant institutional guidelines for the use of secondary survey data.

## Data availability

The data that support the findings of this study are publicly available from the RAND Corporation. The Indonesia Family Life Survey Wave 5 (IFLS-5) public use data can be accessed, following free registration, at https://www.rand.org/well-being/social-and-behavioral-policy/data/ FLS/IFLS/access.html. The analysis code used in this study is available from the corresponding author upon reasonable request.

## Author approval

All authors have read and approved the final version of the manuscript and have agreed to its submission to medRxiv.

## Data Availability

The data supporting the findings of this study are publicly available from the RAND Corporation. The Indonesia Family Life Survey Wave 5 (IFLS-5) public-use data can be accessed following free user registration. The analysis code used in this study is available from the corresponding author upon reasonable request.

https://www.rand.org/well-being/social-and-behavioral-policy/data/FLS/IFLS/access.html

## **A** Data Sources and Variable Construction

This section provides detailed information on data sources and the construction of key variables used in the analysis. It documents the mapping between analytical variables and original IFLS variable names and modules to support reproducibility.

### **A.1** Data Sources

Data were obtained from the fifth wave of the Indonesia Family Life Survey (IFLS-5), a nationally representative household survey conducted in 2014–2015. Disaster-related variables were derived from two survey modules:

- ND1 (`b2_nd1.dta`): records whether the household experienced specific types of natural disasters using multi-select categorical responses.
- ND2 (`b2_nd2.dta`): provides additional quantitative information on disaster events, including frequency, asset loss, and timing.

These modules were linked to the main household and individual datasets using unique household identifiers.

### **A.2** Flood Exposure Variable

Flood exposure (flood_exposure) was defined as a binary household-level indicator. From the ND1 module, households reporting flood exposure were identified using the disaster type coding, where flood events correspond to category “A” within the multi-response field:

- 1 = household experienced flood
- 0 = no reported flood exposure

In cases where ND1 data were unavailable, information from the ND2 module was used as a fallback via a derived indicator (`nd2_is_flood`). The resulting variable was defined at the household level and subsequently assigned to all individuals residing within the same household.

### **A.3** Disaster Risk Score Construction

To capture broader exposure to disaster-related hazards, a composite disaster risk score (`disaster_risk_score`) was constructed reflecting cumulative exposure intensity. It incorporates three components, each standardised using z-score transformation prior to aggregation:

- Disaster frequency (`disaster_frequency`), weight 0.40
- Loss magnitude (`loss_magnitude`), weight 0.40
- Disaster diversity (`disaster_diversity`), weight 0.20

Where components were partially missing, weights were redistributed proportionally across available components to preserve scale consistency. The resulting index provides a continuous measure of household-level disaster burden and was included as a contextual covariate in multilevel models.

### **A.4** Individual-Level Covariate Construction

The wealth index (`wealth_score`) was constructed using principal component analysis (PCA) on log-transformed household consumption (`log_consumption`), household income (`log_income`), and household assets (`log_assets`). Each indicator was aggregated at the household level prior to analysis. Where multiple indicators were available, the first principal component was retained. Where only a single indicator was available, the standardised value of that indicator served as the wealth index directly. The resulting score was subsequently mean-centred and scaled (`wealth_centered`) for inclusion in Bayesian models.

Health-related variables were derived from IFLS health modules. Self-rated health (poor_srh) was defined as a binary indicator corresponding to poor or very poor reported health. Functional limitation (`any_adl_limitation`) was defined as reporting difficulty in at least one activity of daily living (KK03 series). Chronic condition status (`any_chronic_condition`) was defined as the presence of at least one medically diagnosed condition.

Urban–rural classification (`urban_rural`) was derived from the third digit of the IFLS community enumeration area (EA) code, where “1” denotes urban and “2” denotes rural areas.

### **A.5** Variable Harmonisation and Processing

All disaster-related variables were harmonised prior to analysis to ensure consistency across modules. Continuous variables were standardised where appropriate, and derived indicators were constructed to align with the analytical framework. Data processing was implemented in R, with scripts available upon request.

### **A.6** Variable Mapping (IFLS to Analytical Variables)

Tables 6 and 7 provide the complete mapping between analytical variables used in the main text and the original IFLS variable names, source modules, and transformation procedures.

**Table 6:** IFLS variable mapping: source variables and modules (Part A).

| Analytical variable | IFLS variable | Module (file) |
| --- | --- | --- |
| *Outcome* |  |  |
| high_depression | kp items | Health (b3b_kp.dta) |
| theta ($\theta$) | kp items | Health (b3b_kp.dta) |
| *Flood and disaster exposure* |  |  |
| flood_exposure | nd01type (ND1) / nd2_is_flood (ND2) | Disaster (b2_nd1.dta, b2_nd2.dta) |
| disaster_risk_score | nd04, nd07, nd09 | Disaster (b2_nd2.dta) |
| *Demographic and socioeconomic covariates* |  |  |
| age_centered | ar09 | Household roster (bk_ar1.dta) |
| gender | ar07 | Household roster (bk_ar1.dta) |
| marital_status | ar13 | Household roster (bk_ar1.dta) |
| education_level | ar16 | Household roster (bk_ar1.dta) |
| wealth_score/<br>wealth_centered | Multiple | Economic modules (b1_ks*.dta, b3a_hr1/hr2.dta) |
| log_consumption | ks-series items | Consumption modules (b1_ks0–b1_ks4.dta) |
| *Health status covariates* |  |  |
| poor_srh | kk01 | Health (b3b_kk1.dta) |
| any_adl_limitation | kk03 series | Health (b3b_kk3.dta) |
| any_chronic_condition | cd05 | Health (b3b_cd3.dta) |
| *Location* |  |  |
| urban_rural | EA code (digit 3) | Community identifier |
| commid_EA | commid14 | Community identifier |
| *Social trust* |  |  |
| trust_comm_z | tr01–tr30 series | Social capital (b3a_tr.dta) |
| *Resilience indices* |  |  |
| resilience_struct | Derived | Multiple modules |
| resilience_re | Derived (brms) | Primary Bayesian model |

**Table 7:**
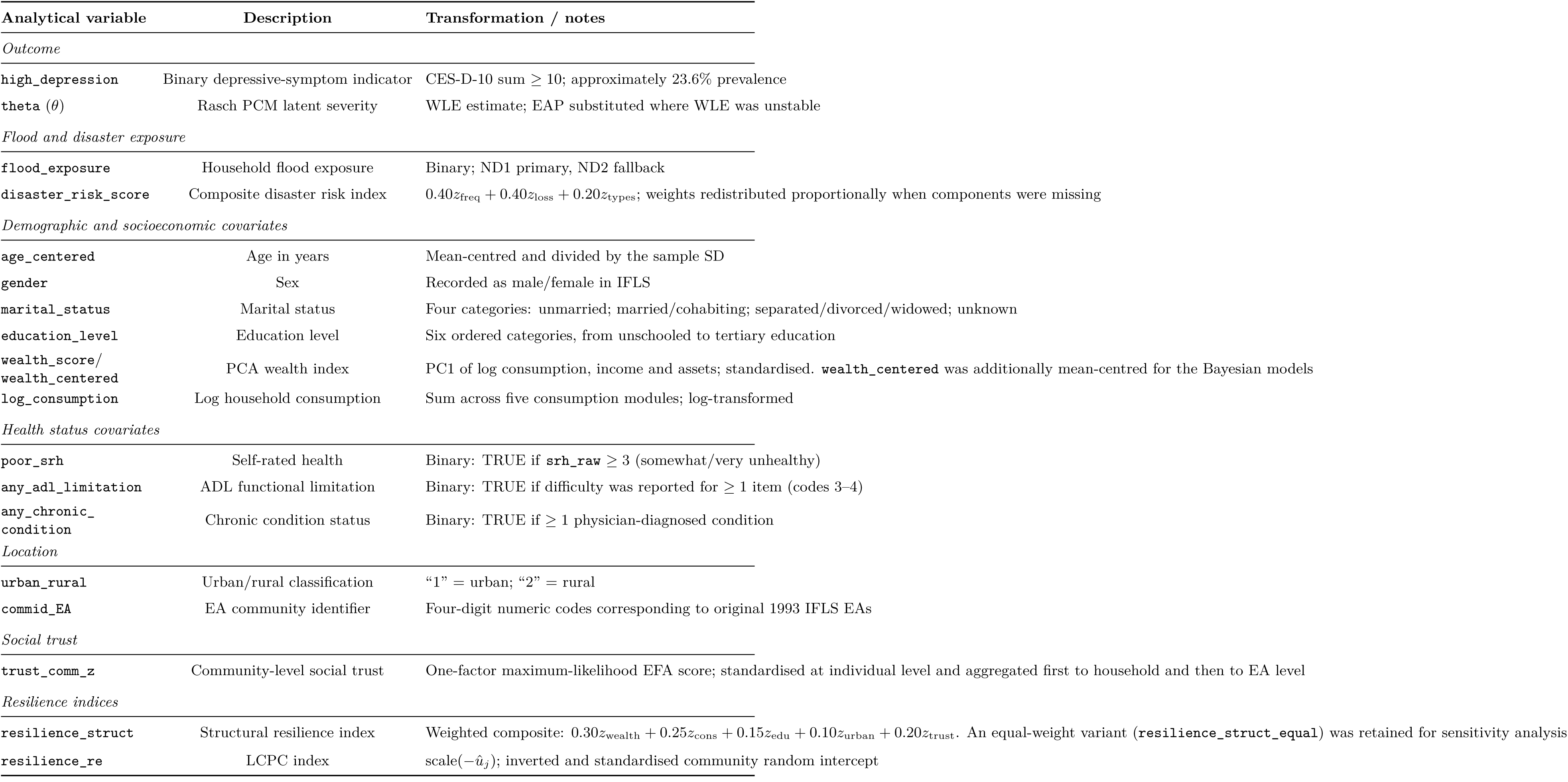
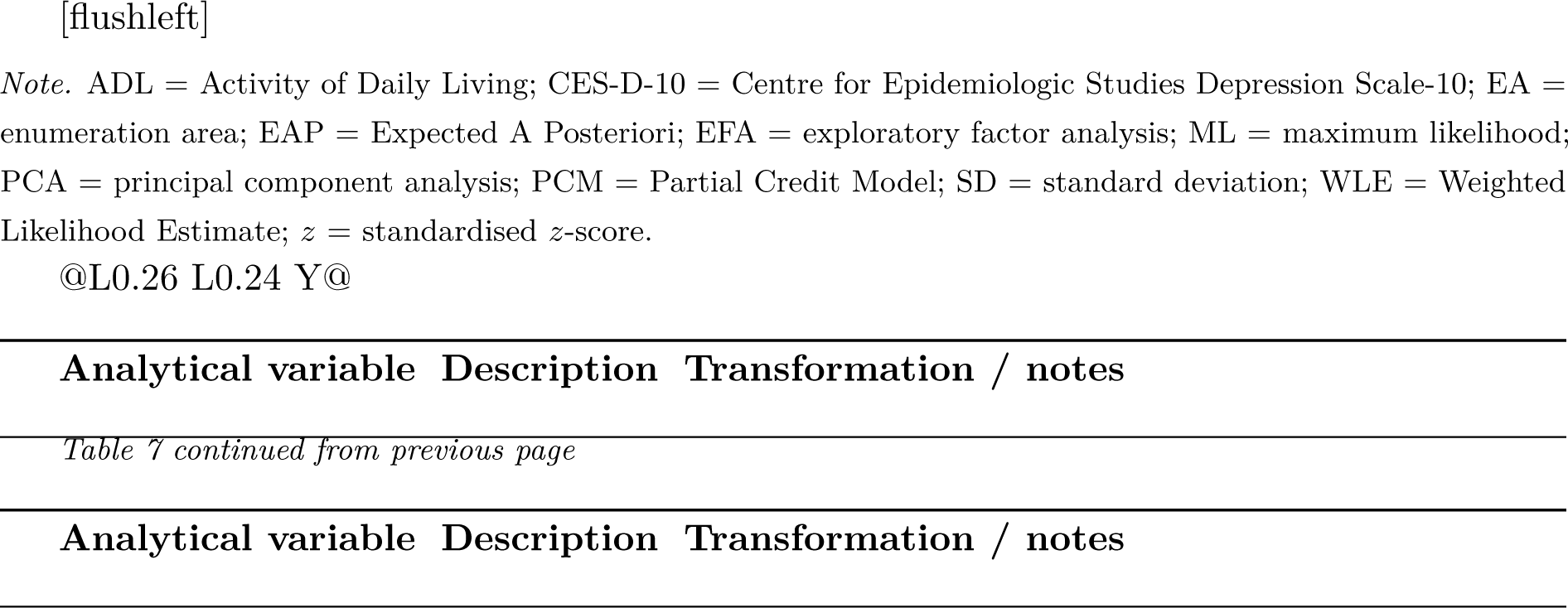
IFLS variable mapping: descriptions and transformations (Part B).

Outcome `high_depression` Binary depression indicator CES-D-10 sum ≥ 10; approx. 23.6% prevalence

`theta` (*θ*) Rasch PCM latent severity WLE estimate; EAP substituted where WLE unstable

Flood and disaster exposure flood_exposure Household flood exposure Binary; ND1 primary, ND2 fallback

`disaster_risk_score` Composite disaster risk index 0.4*z*_freq_ + 0.4*z*_loss_ + 0.2*z*_types_; weights redistributed if components missing

Demographic and socioeconomic covariates age_centered Age in years Mean-centred and divided by SD

`gender` Binary gender Coded male/female

`marital_status` Marital status Four categories: Unmarried; Married/Cohabiting; Sep./Div./Wid.; Unknown

`education_level` Education level Six ordered categories: Unschooled through Tertiary

`wealth_score / wealth_centered` PCA wealth index PC1 of log consumption, income, assets; standardised; `wealth_centered` = further mean-centred for Bayesian models

`log_consumption` Log household consumption Sum across five consumption modules; log-transformed

Health status covariates `poor_srh` Self-rated health Binary: TRUE if srh_raw ≥ 3 (somewhat/very unhealthy)

`any_adl_limitation` ADL functional limitation Binary: TRUE if difficulty with ≥ 1 item (codes 3–4)

`any_chronic_condition` Chronic condition status Binary: TRUE if ≥ 1 physician-diagnosed condition

`Location urban_rural` Urban/rural classification “1” = urban; “2” = rural commid_EA EA community identifier Four-digit numeric codes = original 1993 IFLS EAs

`Social trust trust_comm_z` Community-level social trust One-factor ML EFA score; standardised at individual level; aggregated to household then EA level

Resilience indices `resilience_struct` Structural resilience index Weighted composite: 0.30*z*_wealth_ + 0.25*z*_cons_ + 0.15*z*_edu_ + 0.10*z*_urban_ + 0.20*z*_trust_; equal-weight variant (`resilience_struct_equal`) retained for sensitivity analyses `resilience_re` LCPC index scale(−*u*^*_j_*); inverted standardised community random intercept

### **A.7** Social Trust Variable Construction

Social trust variables were derived from the IFLS social capital module (b3a_tr.dta), including items capturing perceived safety, interpersonal trust, and community attitudes (e.g., tr01–tr07, tr08–tr10, tr23–tr30). All items were harmonised to a common 0–3 scale, with higher values indicating greater trust. Items coded in the direction of distrust (tr06, tr07) were reverse-scored. Respondents with fewer than 50% of items completed were excluded.

A single latent trust factor was estimated using one-factor maximum likelihood exploratory factor analysis using the psych package in R. Individual factor scores were standardised (`trust_z`), then aggregated to the household level (`trust_hh`) and subsequently to the community (enumeration area) level (`trust_comm_z`) by averaging within groups.

### **A.8** Community-Level Aggregation of Indicators

Community-level indicators used in the structural resilience index were derived by aggregating individual- and household-level variables to the enumeration area (EA) level. Specifically, mean household wealth (`wealth_score`), mean log household consumption (`log_consumption`), mean educational attainment (`education_level`), proportion of urban households (`urban_rural`), and mean social trust (`trust_hh`) were computed within each EA. All aggregated indicators were subsequently standardised (z-score transformation) prior to inclusion in the resilience index. Aggregation was performed using all available observations within each EA, with consistency checks applied to ensure alignment between household and individual identifiers.

### **A.9** CES-D-10 Processing and Rasch Calibration Details

In IFLS-5, each CES-D item was collected in long format (module `b3b_kp.dta`) and pivoted to wide form with one column per item type (`kptype`). Items were recoded from the original 1–4 IFLS response scale to the conventional 0–3 metric (0 = rarely or none of the time, 3 = most or all of the time). Two positive-affect items (items E and H: “felt hopeful” and “were happy”) were reverse-scored. Respondents with fewer than seven of the ten items answered were excluded from all analyses.

A Partial Credit Model (PCM) was fitted to the polytomous CES-D-10 item matrix using the R package *eRm* (Mair et al., 2025), which is appropriate for ordered polytomous items under the Rasch paradigm (Masters, 1982). Person ability parameters (*θ*) were estimated as Weighted Likelihood Estimates (WLE) using the R package *TAM* ; where WLE computation failed, Expected A Posteriori (EAP) scores were substituted. The Rasch *θ* scale provides an interval-level measure of latent depressive symptom severity that accounts for differential item difficulty, yielding more robust measurement properties relative to a simple sum score.

The calibrated latent-scale threshold corresponding to the CES-D-10 ≥ 10 cutoff was derived by computing the median *θ* estimate among all respondents with a sum score of exactly 10. The continuous *θ* score was retained for sensitivity analyses examining threshold stability across quantile-defined cut points (60th–85th percentiles of *θ*).

## **B** Community Resilience Index Construction and Validation

### **B.1** Structural Resilience Index Construction

The structural resilience index was constructed to capture the baseline socioeconomic capacity of communities to absorb and recover from flood-related shocks. The index was defined at the enumeration area (EA) level using five aggregated and standardised components derived from household-level data (see appendix section A.7 for variable construction and aggregation procedures):

- Mean household wealth (wealth_score)
- Mean household consumption (log_consumption)
- Mean educational attainment (education_level)
- Proportion of urban households (urban_rural)
- Mean household social trust (trust_hh)

All components were standardised as described in appendix section A prior to index construction.

The structural resilience index was computed as a weighted linear combination:

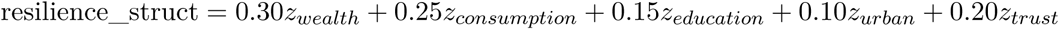

Where one or more components were missing, weights were proportionally redistributed across available components to preserve scale consistency. The resulting index was not re-standardised after aggregation in order to retain interpretability of component contributions.

### **B.2** Weighting Rationale

The weighting structure reflects a theoretically informed prioritisation of economic and material resources as primary determinants of post-disaster recovery capacity. Wealth and consumption were assigned the highest weights (0.30 and 0.25, respectively), reflecting their central role in enabling access to recovery resources, housing stability, and healthcare.

Educational attainment (0.15) serves as a proxy for human capital and preparedness capacity, while urbanisation (0.10) captures access to infrastructure and formal services. Social trust (0.20) represents community cohesion and the capacity for collective action.

This weighting scheme is consistent with the conceptual structure of the Baseline Resilience Indicators for Communities (BRIC) framework (Cutter et al., 2010), which similarly emphasises the dominant role of economic capital while incorporating social and institutional dimensions.

## **C** Sensitivity and Validation Analyses

### **C.1** Threshold Sensitivity Analysis for Depression Classification

Threshold sensitivity analyses across six alternative CES-D *θ* percentile cutpoints (60th–85th) confirmed that classification performance is relatively stable across reasonable threshold choices. CV-AUC ranged from 0.524 (85th percentile, prevalence 15.9%) to 0.573 (60th–65th percentile, prevalence 40.1%), with the primary 80th-percentile cutpoint (prevalence 23.6%) yielding CV-AUC = 0.533 (Figure 8). The monotonic decrease in CV-AUC with increasing threshold stringency reflects the greater difficulty of identifying a smaller, more severely affected minority; the stability of the AUC ordering across thresholds supports the robustness of the comparative conclusions about resilience effects.

**Figure 8:**
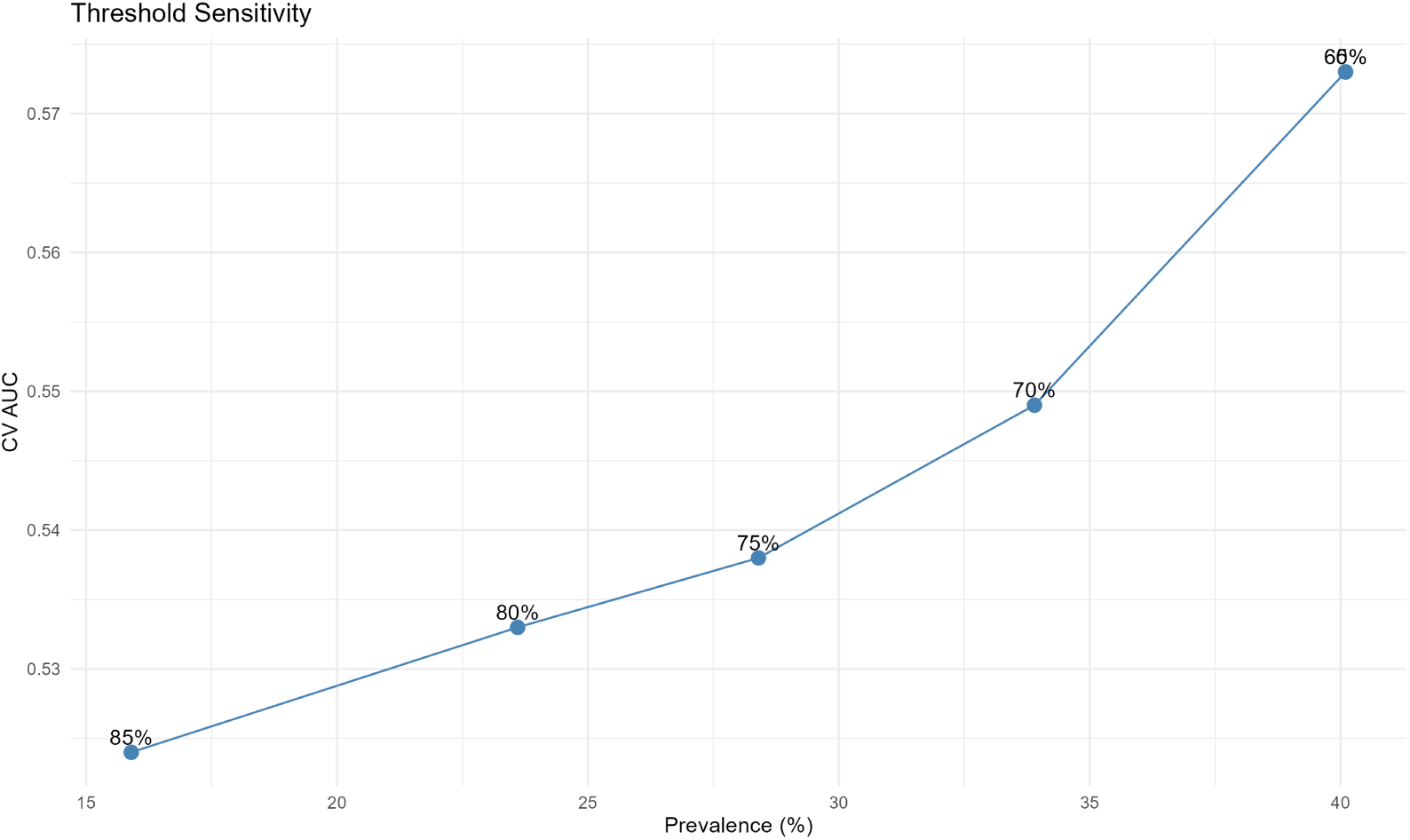
Threshold sensitivity of the binary depression classification. Cross-validated AUC (3-fold, Random Forest) plotted against outcome prevalence across alternative Rasch *θ* percentile cutpoints. Labels indicate the percentile threshold used; the primary *CES* − *D* ≥ 10 classification corresponds to the 80th percentile (*prevalence* ≈ 23.6%). Source: Author’s analysis of IFLS-5 data.

### **C.2** Structural Resilience Index Sensitivity

#### **C.2.1** Equal-Weight Sensitivity Analysis

The primary theory-weighted structural resilience index (component weights: 0.30, 0.25, 0.15, 0.10, 0.20) was compared against an equal-weight alternative assigning a weight of 0.20 to each of the five components. The two indices correlated at *r* = 0.982 across the 312 IFLS original communities (Figure 9), confirming that substantive conclusions are insensitive to the specific weighting scheme within the theoretically motivated range.

**Figure 9:**
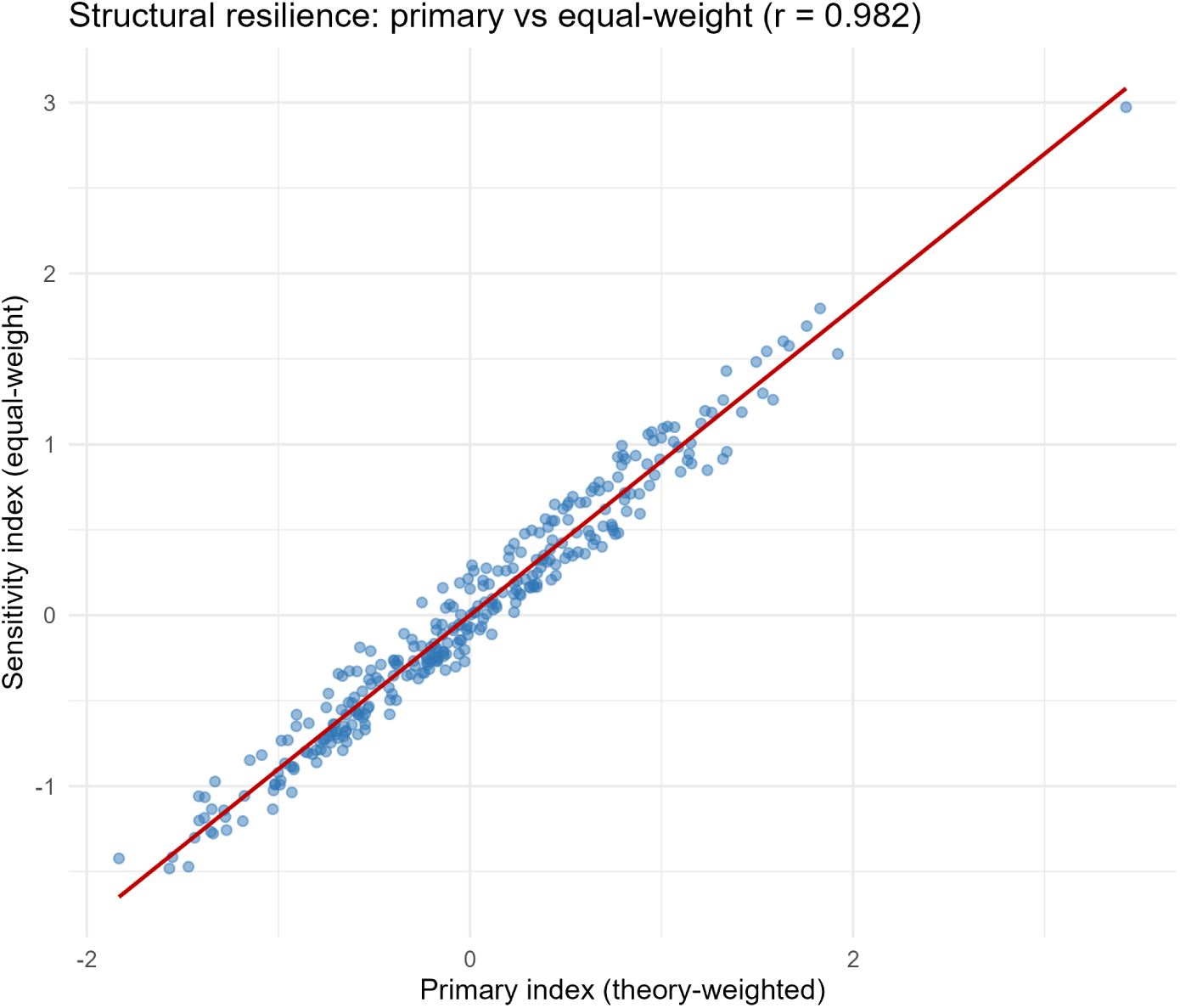
Convergent validity of the structural resilience index: theory-weighted primary index versus equal-weight sensitivity variant across 312 IFLS communities (*r* = 0.982). Source: Author’s analysis of IFLS-5 data.

#### **C.2.2** PCA Convergent Validity

As a further convergent validity check, principal component analysis (PCA) was applied to the five community-level indicator z-scores. The first principal component (PC1) explained 53.5% of the total variance (Figure 10), and the theory-weighted index correlated at *r* = 0.994 with PC1, confirming that the composite captures the dominant axis of between-community socioeconomic differentiation. PC1 loadings were highest for mean consumption (0.564) and mean wealth (0.560), with social trust contributing meaningfully (0.335) and the urban proportion loading substantially more weakly (0.107), consistent with the heterogeneous urbanisation structure of the 1993 IFLS sampling frame (Figure 11).

**Figure 10:**
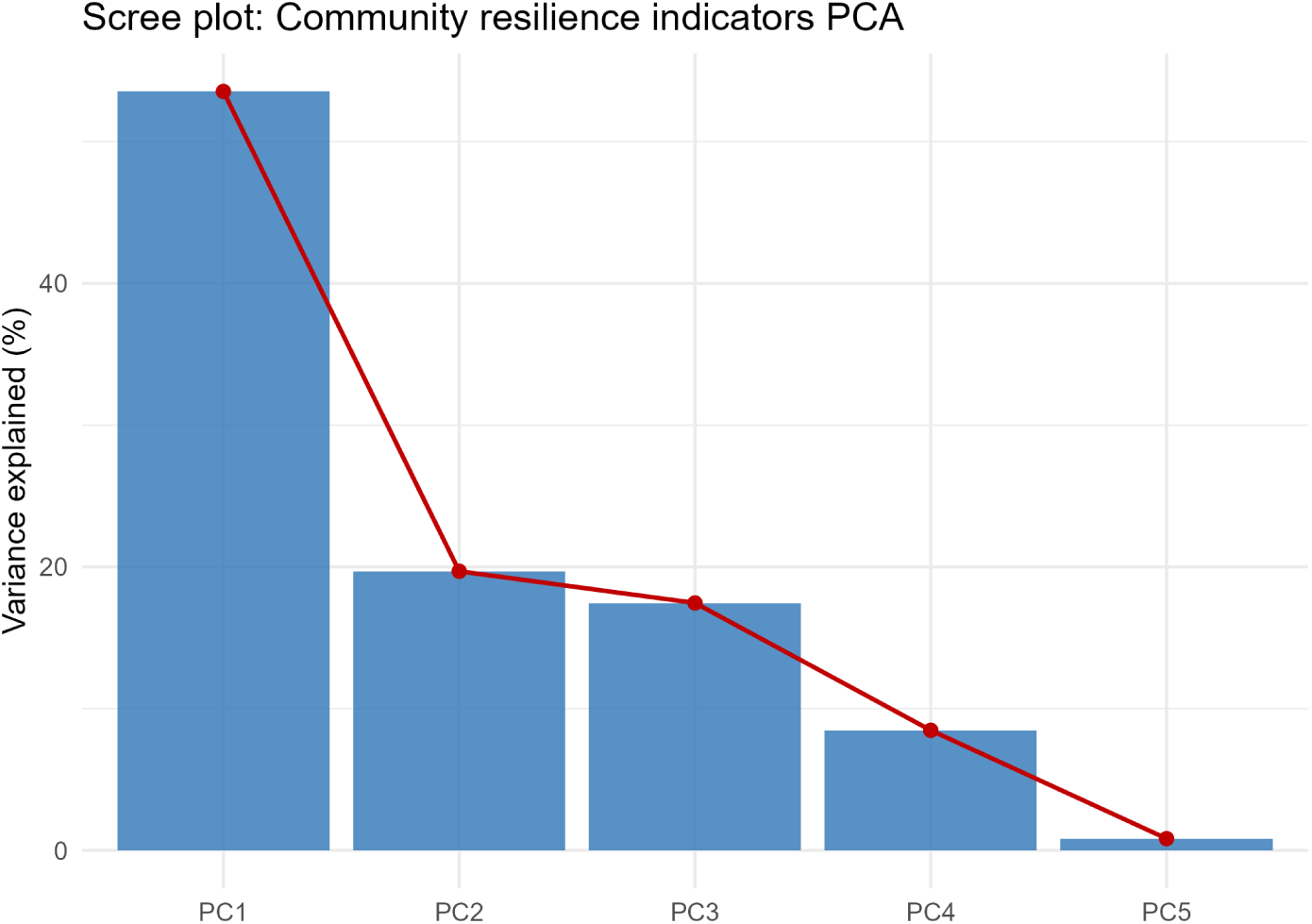
Scree plot for the PCA of the five community resilience indicator components. The first principal component (PC1) explains 53.5% of the total variance, with a pronounced elbow after the first component confirming the unidimensional structure of the indicator space. Source: Author’s analysis of IFLS-5 data.

**Figure 11:**
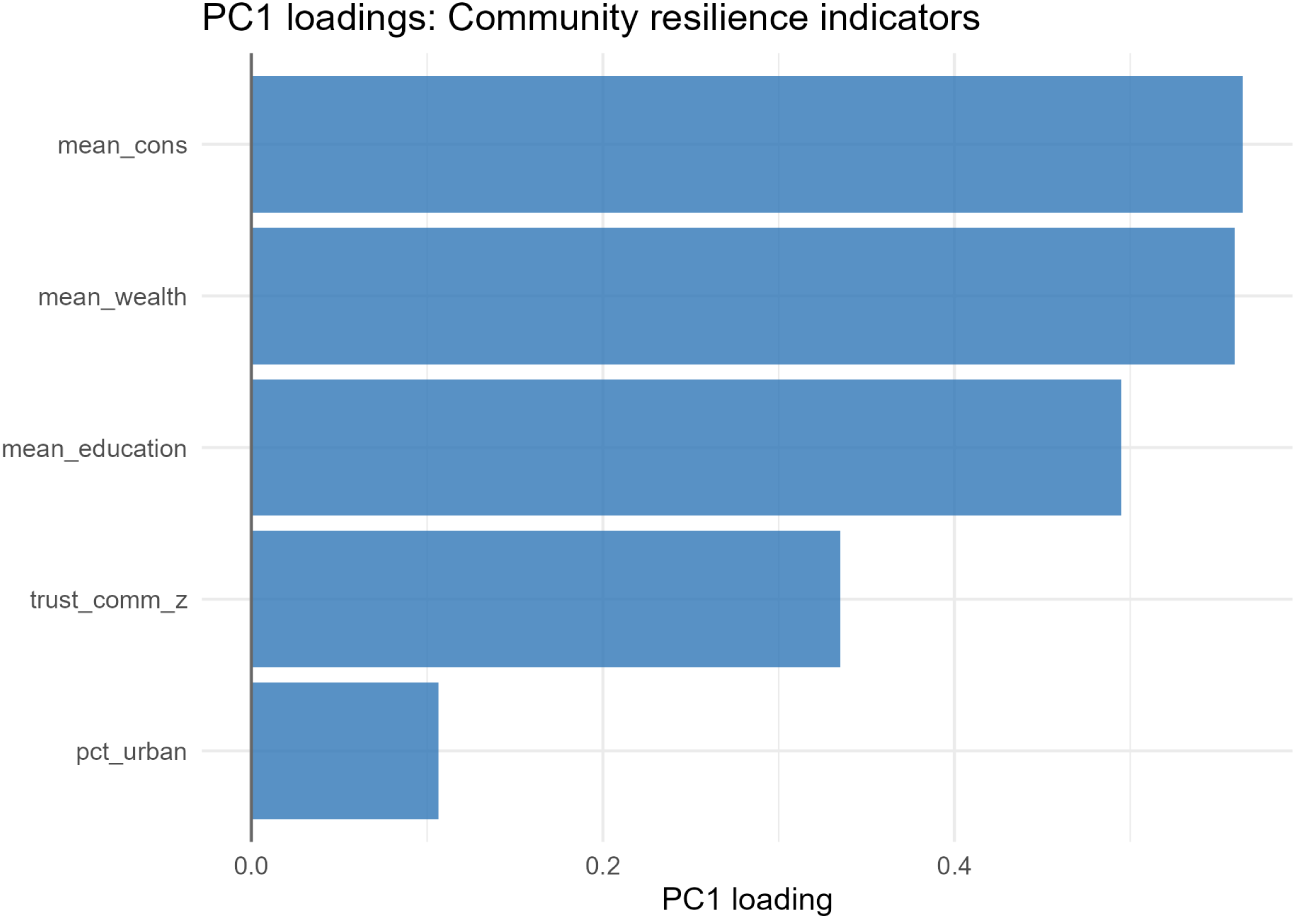
PC1 loadings for the five community resilience indicator components. Mean consumption and mean wealth dominate the first principal component, while the urban proportion loads substantially more weakly, reflecting the heterogeneous urbanisation structure of the 1993 IFLS sampling frame. Source: Author’s analysis of IFLS-5 data.

### **C.3** LCPC Index: Validity and Robustness

The LCPC measure (`resilience_re`) is derived from the posterior mean of the community-level random intercepts of the primary Bayesian model. Its subsequent use as a predictor in the machine learning models introduces a generated regressor concern (Pagan, 1984; Murphy and Topel, 1985): the variable is constructed using the depression outcomes of the same individuals on whom the models are trained, meaning the training data has effectively seen outcome-correlated information when constructing one of its predictors.

#### **C.3.1** Grouped *k*-fold Cross-Validation Procedure

To address this concern for the machine learning predictive models, a grouped *k*-fold cross-validation (GKF) procedure was implemented. The key principle is that each community’s resilience score must be derived from a model that was never trained on that community’s outcomes. To account for potential spatial correlation between neighbouring communities, folds were constructed using province-level blocking rather than random community assignment: all 312 communities were assigned to one of the folds corresponding to the 13 IFLS provinces, ensuring that training communities are always geographically separated from held-out communities. Sensitivity analyses using random community-to-fold assignment (*k* = 5) produced community-level scores correlated at *r* = 0.993 with the province-blocked scores, confirming that substantive conclusions are insensitive ot the fold assignment strategy. The procedure proceeds as follows:

1. All communities within the same province were assigned to the same fold, producing 13 province-level folds. This spatial blocking ensures that training communities are geographically separated from held-out communities, preventing spatial autocorrelation from inflating the apparent out-of-sample performance of the nuisance model.
2. For each fold *f* ∈ {1*, . . .,* 13}:
(a) A logistic regression model was fitted on all individuals from the *k* − 1 remaining folds, using the same fixed-effect predictors as the primary Bayesian model: age (standardised), wealth (standardised), gender, marital status, urban/rural, poor self-rated health, any chronic condition, any ADL limitation, and disaster risk score.
(b) Fixed-effects-only predicted probabilities 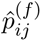 were generated for all individuals *i* in the held-out communities *j* ∈ fold *f* .
(c) Individual-level residuals were computed as 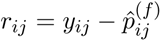, where *y_ij_*is the binary depression outcome.
(d) Community-level mean residuals were computed: 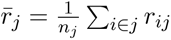.
3. After all five folds, each of the 312 communities has an out-of-fold mean residual *r̂_j_*. These were sign-inverted and standardised to produce `resilience_re_gkf`:

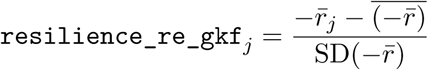

Higher values indicate communities whose observed depression rates were lower than predicted by the fixed-effect model, consistent with the interpretation of `resilience_re`.

The GKF scores (`resilience_re_gkf`) are used exclusively in the machine learning hybrid models. The full `resilience_re` derived from Bayesian random intercepts is retained for the moderation and variance decomposition analyses, where it serves as a community-level contextual construct rather than an individual-level predictor.

#### **C.3.2** Limitations of the Approximation

The GKF procedure uses a logistic regression nuisance model rather than the full Bayesian multilevel model, meaning the predicted probabilities do not account for community-level clustering. The community mean residuals therefore absorb both genuine community-level protective capacity and noise from the nuisance model’s inability to capture community random effects. This is an inherent limitation of the approximation; a fully correct solution would require leave-community-out refitting of the Bayesian model, which is computationally prohibitive with 312 communities. For the Bayesian moderation analysis, the full random-intercept-based construct is retained and full propagation of uncertainty from the primary model to the moderation model was not implemented. Those estimates should therefore be interpreted as indicative rather than fully uncertainty-quantified, as noted in the limitations (Section 6 of the main text).

### **C.4** Software and Implementation Details

All analyses were conducted in R (version ≥ 4.4). Bayesian multilevel models were fitted using the brms package (version ≥ 2.18), with Stan as the computational backend. Machine learning analyses were implemented using the randomForest (version 4.7) and xgboost (version 1.7) packages.

Data preprocessing, model fitting, and result extraction were performed using custom R scripts. The full analytic pipeline is available from the corresponding author upon reasonable request.

### **C.5** Summary of Analytic Variables

This section contains an overview of the analytical variables used in this research. They are shown in Table 8

**Table 8:**
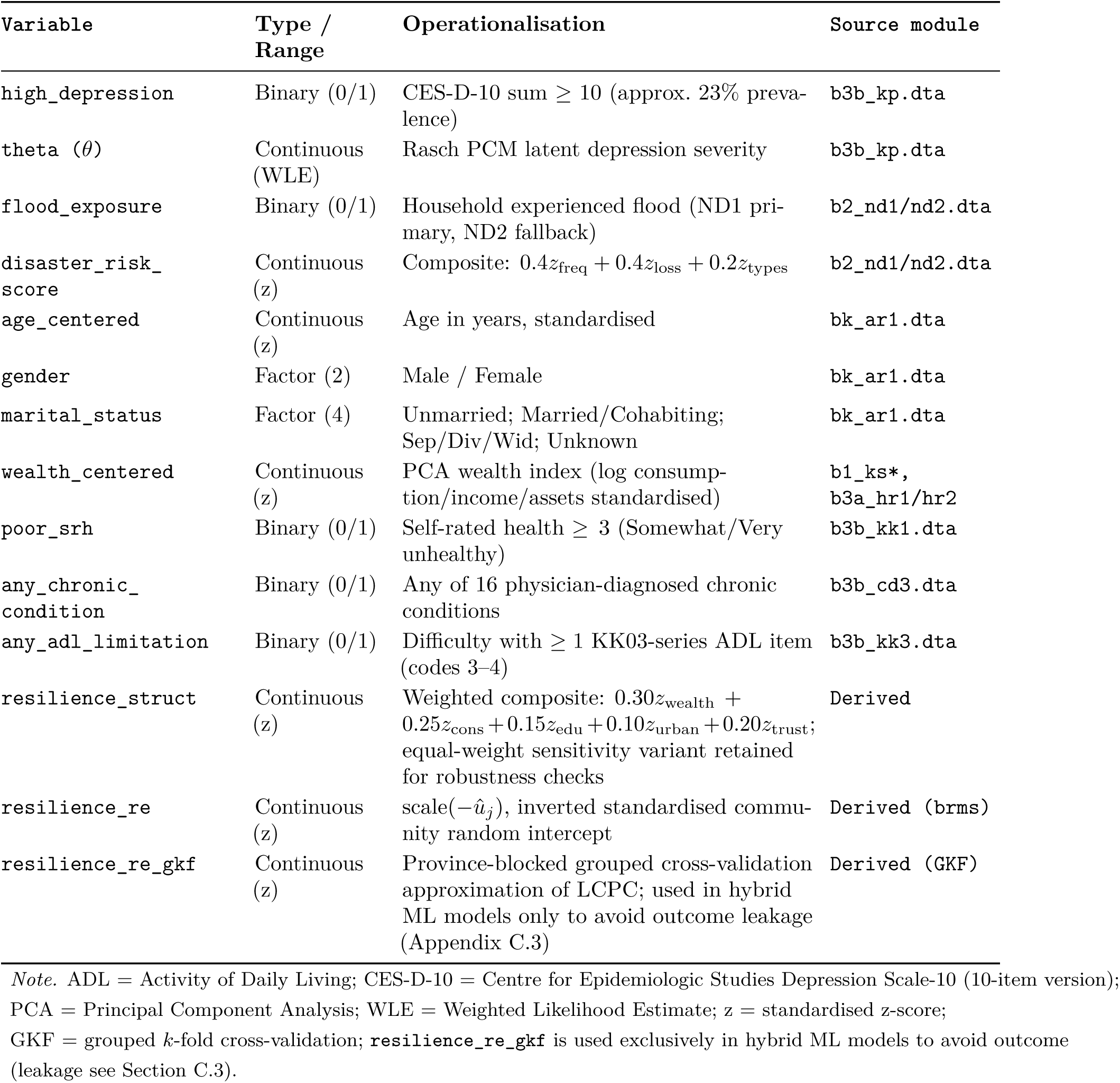
Summary of analytic variables, definitions, and source modules.

| Variable | Type / Range | Operationalisation | Source module |
| --- | --- | --- | --- |
| high_depression | Binary (0/1) | CES-D-10 sum $\geq 10$ (approx. 23% prevalence) | b3b_kp.dta |
| theta ($\theta$) | Continuous (WLE) | Rasch PCM latent depression severity | b3b_kp.dta |
| flood_exposure | Binary (0/1) | Household experienced flood (ND1 primary, ND2 fallback) | b2_nd1/nd2.dta |
| disaster_risk_score | Continuous (z) | Composite: $0.4z_{\text{freq}} + 0.4z_{\text{loss}} + 0.2z_{\text{types}}$ | b2_nd1/nd2.dta |
| age_centered | Continuous (z) | Age in years, standardised | bk_ar1.dta |
| gender | Factor (2) | Male / Female | bk_ar1.dta |
| marital_status | Factor (4) | Unmarried; Married/Cohabiting; Sep/Div/Wid; Unknown | bk_ar1.dta |
| wealth_centered | Continuous (z) | PCA wealth index (log consumption/income/assets standardised) | b1_ks*,<br>b3a_hr1/hr2 |
| poor_srh | Binary (0/1) | Self-rated health $\geq 3$ (Somewhat/Very unhealthy) | b3b_kk1.dta |
| any_chronic_condition | Binary (0/1) | Any of 16 physician-diagnosed chronic conditions | b3b_cd3.dta |
| any_adl_limitation | Binary (0/1) | Difficulty with $\geq 1$ KK03-series ADL item (codes 3–4) | b3b_kk3.dta |
| resilience_struct | Continuous (z) | Weighted composite: $0.30z_{\text{wealth}} + 0.25z_{\text{cons}} + 0.15z_{\text{edu}} + 0.10z_{\text{urban}} + 0.20z_{\text{trust}}$; equal-weight sensitivity variant retained for robustness checks | Derived |
| resilience_re | Continuous (z) | scale($-\hat{u}_j$), inverted standardised community random intercept | Derived (brms) |
| resilience_re_gkf | Continuous (z) | Province-blocked grouped cross-validation approximation of LCPC; used in hybrid ML models only to avoid outcome leakage (Appendix C.3) | Derived (GKF) |

*Note.* ADL = Activity of Daily Living; CES-D-10 = Centre for Epidemiologic Studies Depression Scale-10 (10-item version); PCA = Principal Component Analysis; WLE = Weighted Likelihood Estimate; z = standardised z-score; GKF = grouped $k$ -fold cross-validation; **resilience\_re\_gkf** is used exclusively in hybrid ML models to avoid outcome (leakage see Section C.3).

## **D** Additional Model Specifications

### **D.1** Variance Decomposition Model Specifications

The nested sequence of Bayesian multilevel logistic regression models fitted for variance decomposition is as follows:

**M0** (Null model):

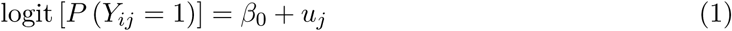

**M1** (+ individual covariates):

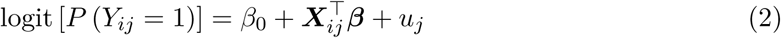

where ***X****_ij_*includes age (centred), gender, marital status, wealth (centred), urban/rural classification, poor self-rated health, any chronic condition, and any ADL limitation.

**M2** (+ disaster exposure):

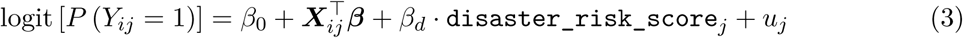

**M3** (+ structural resilience):

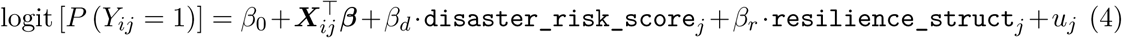

In all models *u_j_* ∼ N (0*, σ_u_*^2^) is the community-level random intercept. The intraclass correlation coefficient (ICC) on the latent logistic scale is computed as:

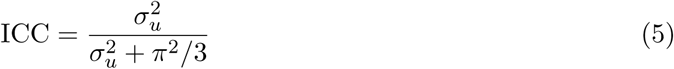

The proportional change in variance (PCV) at each step is computed relative to the empty model M0:

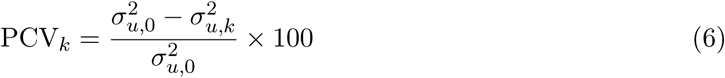

where *σ_u,0_*^2^ denotes the between-community variance from M0 and *σ_u,k_*^2^ denotes the between-community variance from model *k*. Credible intervals for PCV were obtained by propagating uncertainty from the posterior distributions of *σ_u_*^2^ across model comparisons.

### **D.2** Resilience Moderation Model Specification

The structural moderation model takes the form:

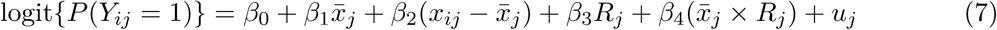

where *x̂_j_* is the community-level proportion of flood-exposed households (between-community component), *x_ij_* − *x̂_j_* is the individual deviation from that mean (within-community component), and *R_j_* denotes the standardised resilience score for community *j*. The statistical moderation model has an identical structure with LCPC substituted for structural resilience.

The interaction coefficient *β*_4_ is the parameter of primary interest: a negative value indicates that communities with higher resilience exhibit a weaker positive association between aggregate flood prevalence and individual depression risk, consistent with the buffering hypothesis. Simple slopes for the moderation effect were computed from the posterior distributions of *β*_1_ and *β*_4_, yielding the estimated effect of flood exposure on depression log-odds at resilience values of −1, 0, and +1 SD. All continuous predictors were standardised prior to model fitting. Conditional effects were evaluated using posterior predictive draws (*n* = 800).

### **D.3** Individual-Level SHAP Detail

Figures 12–14 provide the individual-level SHAP diagnostics summarised in the main text. The waterfall plot (Figure 12) decomposes the prediction for a representative high-risk individual; the dependence plots (Figures 13 and 14) contrast the systematic negative association of LCPC with the diffuse, near-zero contribution of structural resilience across the full range of each index.

**Figure 12:**
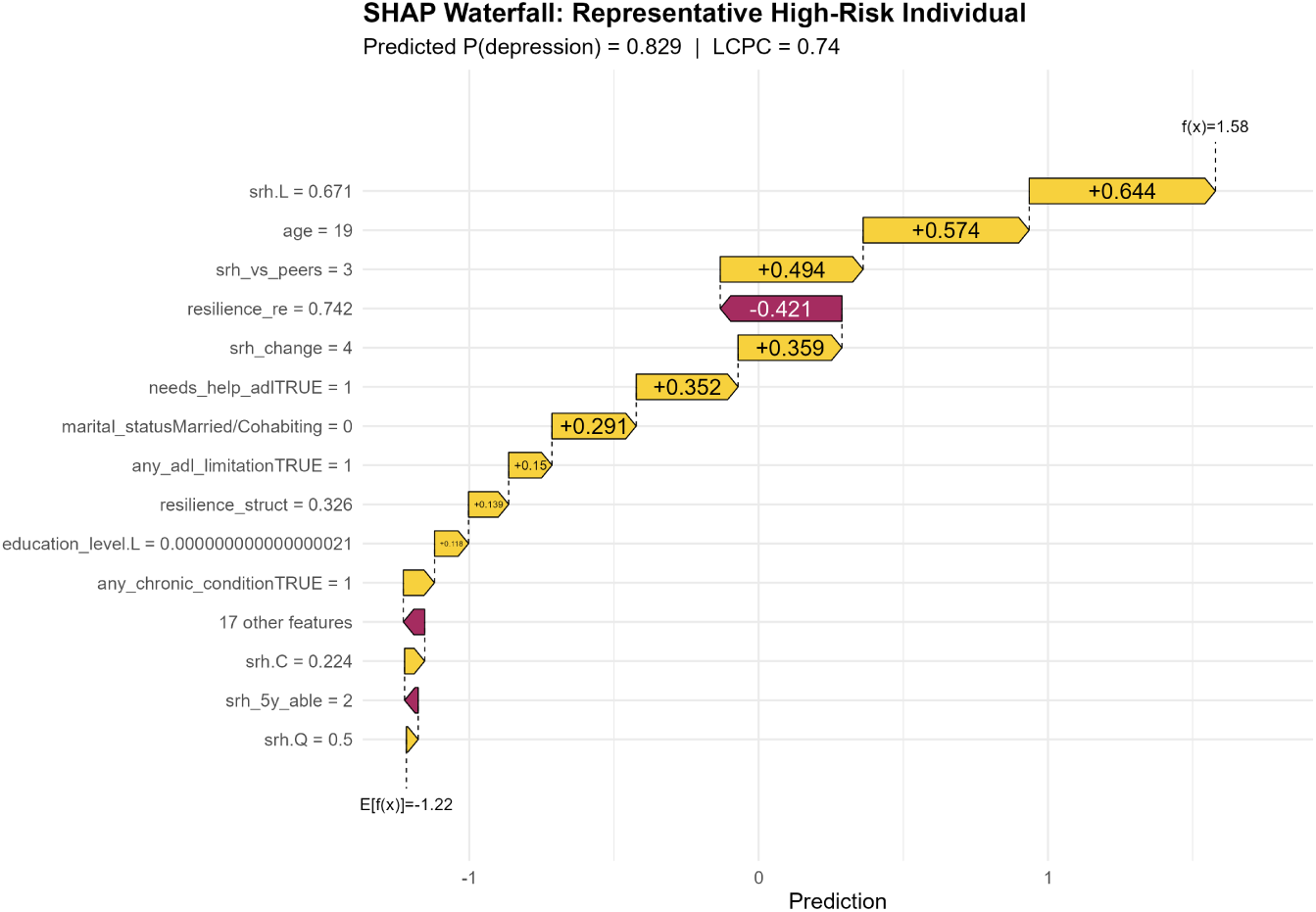
SHAP waterfall plot for a representative high-risk individual (predicted *P* (depression) = 0.829; `resilience_re` = 0.74). Each bar shows the contribution of one feature to the deviation from the expected model output. Despite multiple health and sociodemographic risk factors pushing the prediction upward, LCPC exerts a substantial downward correction (−0.421), illustrating the protective role of community context at the individual level. Source: Author’s analysis of IFLS-5 data.

**Figure 13:**
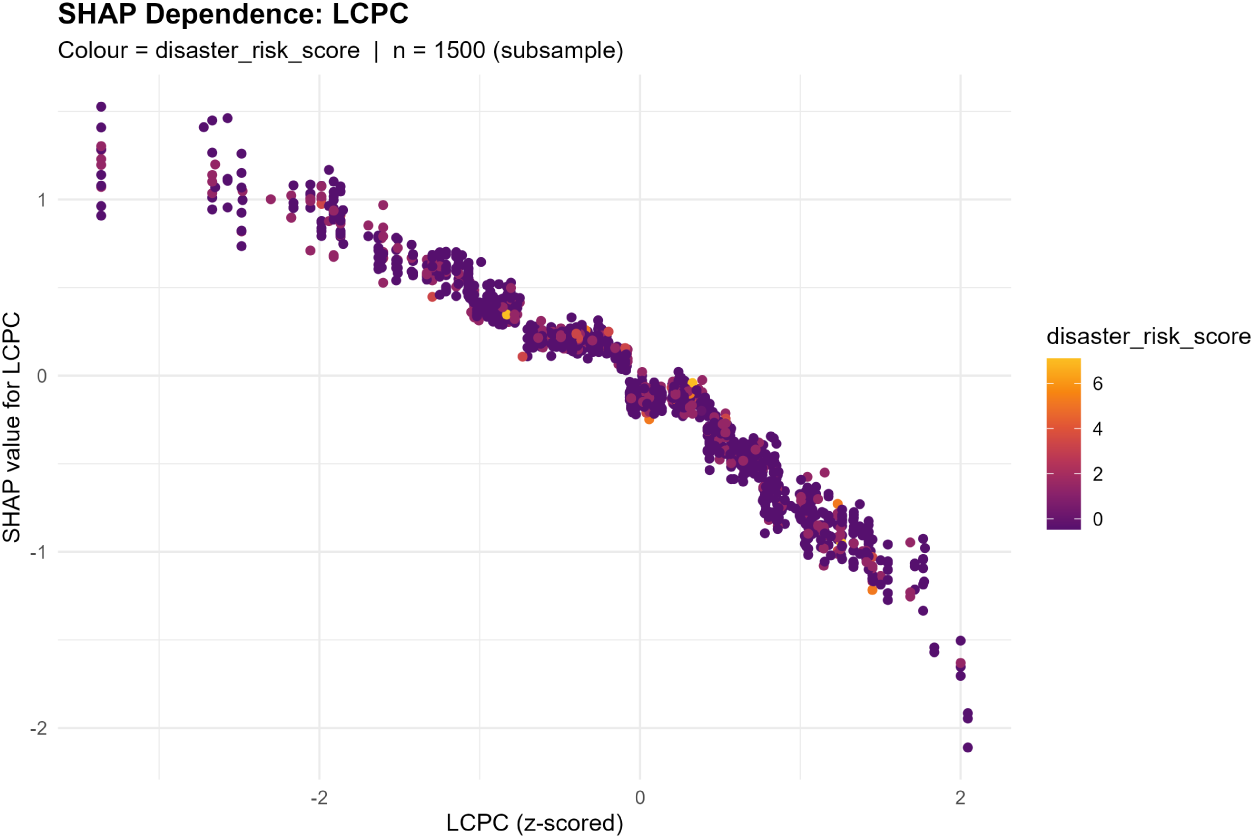
SHAP dependence plot for `resilience_re` (LCPC) in the XGBoost Hybrid-Both model (n = 1,500 subsample). The monotonically negative slope confirms that individuals in higher-resilience communities receive systematically lower depression-risk contributions from this feature. Point colour encodes the disaster risk score. Source: Author’s analysis of IFLS-5 data.

**Figure 14:**
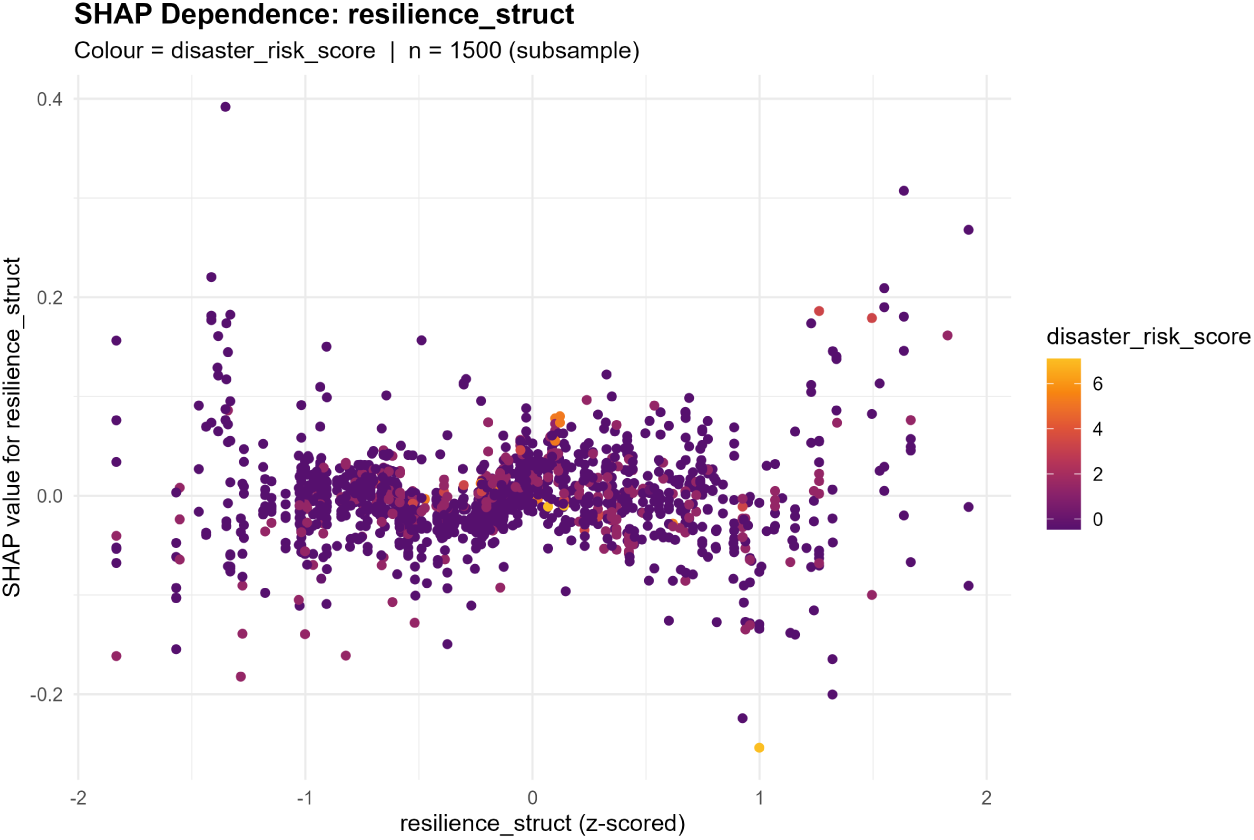
SHAP dependence plot for `resilience_struct` in the XGBoost Hybrid-Both model (n = 1,500 subsample). Unlike LCPC, structural resilience contributions are diffuse and centred near zero across the full range, with no consistent monotonic trend, consistent with its smaller share of overall feature importance. Source: Author’s analysis of IFLS-5 data.

